# Distinct brain atrophy progression subtypes underlie phenoconversion in isolated REM sleep behaviour disorder

**DOI:** 10.1101/2024.09.05.24313131

**Authors:** Stephen Joza, Aline Delva, Christina Tremblay, Andrew Vo, Marie Filiatrault, Max Tweedale, John-Paul Taylor, John T. O’Brien, Michael Firbank, Alan Thomas, Paul C. Donaghy, Johannes Klein, Michele Hu, Petr Dusek, Stanislav Marecek, Zsoka Varga, Stephane Lehericy, Isabelle Arnulf, Marie Vidailhet, Jean-Christophe Corvol, Jean-François Gagnon, Ronald B. Postuma, Alain Dagher, Richard Camicioli, Howard Chertkow, Simon Lewis, Elie Matar, Kaylena A. Ehgoetz Martens, Lachlan Churchill, Michael Sommerauer, Sinah Röttgen, Per Borghammer, Karoline Knudsen, Allan K. Hansen, Dario Arnaldi, Beatrice Orso, Pietro Mattioli, Luca Roccatagliata, Oury Monchi, Shady Rahayel

**Affiliations:** The Neuro (Montreal Neurological Institute-Hospital), McGill University, Montreal H3A 2B4, Canada; Centre for Advanced Research in Sleep Medicine, CIUSSS-NÎM – Hôpital du Sacré-Cœur de Montréal, Montreal H4J 1C5, Canada; Translational and Clinical Research Institute, Newcastle University, Newcastle, UK; Department of Psychiatry, University of Cambridge School of Clinical Medicine, Cambridge, UK; Oxford Parkinson’s Disease Centre and Division of Neurology, Nuffield Department of Clinical Neurosciences, University of Oxford, Oxford, UK; Department of Neurology and Centre of Clinical Neurosciences, First Faculty of Medicine, Charles University and General University Hospital, Prague, Czechia; Institut du Cerveau – Paris Brain Institute – ICM, Sorbonne Université, INSERM UMR1127, CNRS 7225, Clinical Investigation Centre (CIC) Paris 75013, France; Department of Psychology, Université du Québec à Montréal, Montreal H2X 3P2, Canada; Research Centre, Institut universitaire de gériatrie de Montréal, Montreal H3W 1W5, Canada; Department of Neurology, Montreal General Hospital, Montreal H3G 1A4, Canada; Division of Neurology, Department of Medicine, and Neuroscience and Mental Health Institute, University of Alberta, Edmonton, Canada; Department of Medicine (Neurology), University of Toronto, Toronto, Ontario, Canada; Rotman Research Institute, Baycrest Health Services, Toronto, Ontario, Canada; Parkinson’s Disease Research Clinic, Macquarie Medical School, Macquarie University, Sydney, Australia; Parkinson’s Disease Research Clinic, Brain and Mind Centre, University of Sydney, Camperdown NSW 2050, Australia; Department of Kinesiology and Health Sciences, University of Waterloo, Waterloo N2L 3G1, Canada; Centre of Neurology, Department of Parkinson, Sleep and Movement Disorders, University Hospital Bonn, Bonn, Germany; Department of Neurology, University Hospital Cologne, Faculty of Medicine, University of Cologne, Cologne, Germany; Institute of Neuroscience and Medicine (INM-3), Forschungszentrum Jülich, Jülich, Germany; Department of Nuclear Medicine and PET, Aarhus University Hospital, Aarhus DK-8200, Denmark; Department of Neuroscience, Rehabilitation, Ophthalmology, Genetics, Maternal and Child Health (DINOGMI), Clinical Neurology, University of Genoa, 16132 Genoa, Italy; IRCCS Ospedale Policlinico San Martino, 16132 Genoa, Italy; Department of Radiology, Radio-Oncology, and Nuclear Medicine, University of Montreal, Montreal H3T 1A4, Canada; Department of Medicine, University of Montreal, Montreal H3T 1A4, Canada

**Keywords:** REM sleep behaviour disorder, Parkinson’s disease, dementia with Lewy bodies, MRI, subtyping, machine learning

## Abstract

**Background:** Synucleinopathies manifest as a spectrum of disorders that vary in features and severity, including idiopathic/isolated REM sleep behaviour disorder (iRBD) and dementia with Lewy bodies. Patterns of brain atrophy in iRBD are already reminiscent of what is later seen in overt disease and are related to cognitive impairment, being associated with the development of dementia with Lewy bodies. However, how brain atrophy begins and progresses remains unclear.

**Methods:** A multicentric cohort of 1,134 participants, including 538 patients with synucleinopathies (451 with polysomnography-confirmed iRBD and 87 with dementia with Lewy bodies) and 596 healthy controls, was recruited from 11 international study centres and underwent T1-weighted MRI imaging and longitudinal clinical assessment. Scans underwent vertex-based cortical surface reconstruction and volumetric segmentation to quantify brain atrophy, followed by parcellation, ComBAT scan harmonization, and piecewise linear z-scoring for age and sex. We applied the unsupervised machine learning algorithm, Subtype and Stage Inference (SuStaIn), to reconstruct spatiotemporal patterns of brain atrophy progression and correlated the distinct subtypes with clinical markers of disease progression.

**Results:** SuStaIn identified two unique subtypes of brain atrophy progression: 1) a “cortical-first” progression subtype characterized by atrophy beginning in the frontal lobes followed by the temporal and parietal areas and remaining cortical areas, with the involvement of subcortical structures at later stages; and 2) a “subcortical-first” progression subtype, which involved atrophy beginning in the limbic areas, then basal ganglia, and only involving cortical structures at late stages. Patients classified to either subtype had higher motor and cognitive disease burden and were more likely to phenoconvert to overt disease compared with those that were not classifiable. Of the 84 iRBD patients who developed overt disease during follow-up, those with a subcortical-first pattern of atrophy were more likely to phenoconvert at earlier SuStaIn stages, particularly to a parkinsonism phenotype. Conversely, later disease stages in both subtypes were associated with more imminent phenoconversion to a dementia phenotype.

**Conclusions:** Patients with synucleinopathy can be classified into distinct patterns of atrophy that correlate with disease burden. This demonstrates insights into underlying disease biology and the potential value of categorizing patients in clinical trials.

## Background

Synucleinopathies are pathologically defined by the misfolding and aggregation of alpha-synuclein [1]. During the prodromal phases of disease, patients manifest a variety of deficits in multiple clinical domains, including cognitive and motor abnormalities, olfactory dysfunction, constipation, dysautonomia, and sleep disorders [2]. One highly studied prodromal phenotype is idiopathic/isolated REM sleep behaviour disorder (iRBD), a parasomnia characterized by dream enactment behaviours during REM sleep [3]. The vast majority of patients with iRBD will eventually develop an overt and clinically-defined disorder, mainly dementia with Lewy bodies and Parkinson’s disease, and less frequently multiple system atrophy [4].

As a prodromal synucleinopathy, clinical changes and patterns of brain atrophy in iRBD are already reminiscent of what is seen in overt disease [4–6]. In particular, iRBD patients with concomitant mild cognitive impairment have more extensive cortical and subcortical abnormalities compared to those without mild cognitive impairment, with the severity of atrophy predicting subsequent development of dementia with Lewy bodies [7–9]. This supports the notion that substantial variability exists between iRBD patients during this prodromal phase, with some destined to develop dementia earlier in their disease course than others [10]. *In silico* modelling of atrophy in iRBD, based on computational spreading models of alpha-synuclein [11–13], has demonstrated that gene expression and structural connectivity jointly influence brain neurodegeneration [5]. Notably, a closer match between the *in silico* atrophy pattern and the patient’s actual atrophy pattern correlates with increased cognitive impairment but not motor impairment in iRBD [5]. Identifying patterns in this variability might be useful for prognostic purposes and allow more precise selection of patients for future therapeutic trials [3]. However, the changes in brain morphology that begin during iRBD and eventually progress toward the development of dementia remain unclear.

To better understand the relationships between interindividual variability within iRBD patients and their subsequent transition to dementia and parkinsonism, a systematic investigation of the specific sequential brain changes leading to dementia with Lewy bodies is needed. Several studies have documented the longitudinal brain changes taking place over time in iRBD [9,14,15], but these have been restricted by a limited number of patients, the high level of inter-assessment variability in imaging techniques, and the extended follow-up delay between the diagnosis of iRBD and phenoconversion.

In this study, we performed a comprehensive quantification of brain atrophy in iRBD and dementia with Lewy bodies and reconstructed the subtypes of spatiotemporal changes in brain atrophy progression from cross-sectional data to understand their associations with clinical disease progression. We compiled the largest collection of structural brain MRI data acquired to date in patients with iRBD (n=451 from 11 international study centres). To derive atrophy-driven subtypes of iRBD and their associated patterns of progression, we performed vertex-based cortical surface analysis of thickness and subcortical volume quantification on the complete dataset and applied the Subtype and Stage Inference (SuStaIn) model, an unsupervised machine learning algorithm that uses a combined disease progression modelling and clustering approach on cross-sectional scans of patients at different stages of a clinical continuum [16]. Finally, we describe the clinical characteristics and phenoconversion status of the resulting data-driven subtypes to gain an understanding of the relationship between patterns of atrophy in iRBD and the development of dementia and parkinsonism.

## Methods

### Participants

A total of 1134 participants were recruited for this study and underwent T1-weighted brain MRI imaging and clinical assessments (see Figure 1 for an overview of the study protocol). Of these, 451 had polysomnography-confirmed iRBD, 87 had dementia with Lewy bodies, and 596 were healthy controls recruited in every centre.

**Figure 1:**
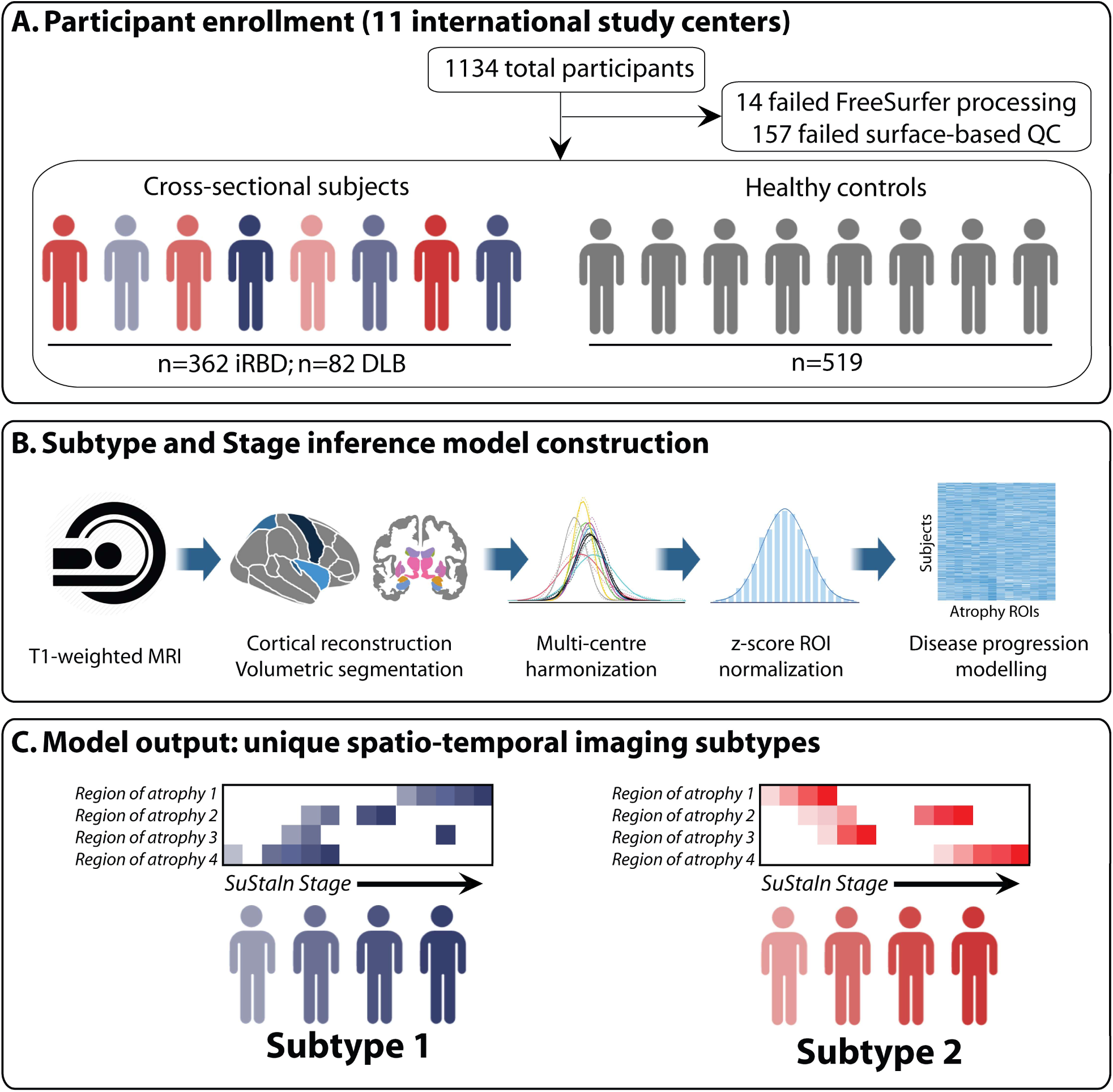
Overview of study design, data processing, and subtype modelling. **(A)** 1,134 participants were recruited, with 171 participants excluded after quality control. **(B)** T1-weighted MRI scans underwent cortical reconstruction, volumetric segmentation, multi-centre harmonization, z-score normalization, and disease progression modelling using the SuStaIn algorithm. **(C)** SuStaIn identified two distinct subtypes of brain atrophy progression, each with unique spatiotemporal patterns and stages of disease. DLB = dementia with Lewy bodies; iRBD = idiopathic/isolated REM sleep behaviour disorder; QC = quality control; ROI = region of interest.

Participant recruitment by study centre and disease group are detailed in Supplementary Table 1. Participants were recruited from Newcastle University (n=188), the Oxford Parkinson’s Disease Centre (n=147), the Department of Neurology at Charles University (n=140), the Movement Disorders clinic at the Hôpital de la Pitié-Salpêtrière (n=130), the Centre for Advanced Research on Sleep Medicine at the Hôpital du Sacré-Cœur de Montréal (n=125), the COMPASS-ND Study from the Canadian Consortium on Neurodegeneration in Aging (CCNA; n=71)[17–19], the Parkinson’s Disease Research Clinic at the University of Sydney (n=56), the Department of Neurology at the University of Cologne (n=47), Aarhus University Hospital (n=38), the IRCCS Ospedale Policlinico San Martino in Genoa (n=29), as well as part of the Parkinson’s Progression Markers Initiative (PPMI; n=305)[20]. A subset of the iRBD patients (n=182, 40%) included in this study were part of previous studies investigating prodromal atrophy in synucleinopathies [5,6].

Patients with iRBD were diagnosed using the International Classification of Sleep Disorders, third edition diagnostic criteria [21], including video-polysomnography, and underwent clinical assessments to confirm absence of dementia with Lewy bodies, Parkinson’s disease, and multiple system atrophy at the closest examination in time to the MRI acquisition. Patients with iRBD were followed longitudinally approximately every 6-12 months in every centre to assess for the development of dementia and parkinsonism (phenoconversion). Clinical assessments used at all sites included cognitive testing with either the Montreal Cognitive Assessment (MoCA) or the Mini-Mental State Examination (MMSE), and motor examination using the Movement Disorders Society – Unified Parkinson’s Disease Rating Scale, part III (MDS-UPDRS-III) or the original 1987 version (UPDRS-III). Patients with probable dementia with Lewy bodies were diagnosed using previously published criteria [22]. The iRBD and dementia with Lewy bodies patients were used for the main modelling findings of this study since previous studies have shown that brain atrophy in iRBD is strongly associated with cognitive impairment and predicts dementia with Lewy bodies compared to Parkinson’s disease [5,7,8]. Including dementia with Lewy bodies therefore provided atrophy measurements from the later stages of brain disease progression when iRBD converts to dementia with Lewy bodies. However, to ensure that this did not drive the subtyping in iRBD, we repeated the analyses using a sample of 142 Parkinson’s disease patients with probable RBD. These patients were recruited from the PPMI baseline cohort, and the presence of probable RBD was defined by a cut-off score ≥5 on the RBD Screening Questionnaire [23].

Ethics approval was obtained from the local institutional boards of each centre with subject consent in accordance with the Declaration of Helsinki. The current study was approved by the Research Ethics Board of the McGill University Health Centre and the Centre intégré universitaire de santé et de services sociaux du Nord-de-l’Île-de-Montréal.

### MRI acquisition and processing

Structural T1-weighted brain MRI scans were acquired at each site and are detailed in Supplementary Table 1. T1-weighted scans underwent cortical reconstruction and volumetric segmentation using the FreeSurfer 7.1.1 image analysis suite (http://surfer.nmr.mgh.harvard.edu). The technical details of the FreeSurfer procedure have been described previously [5]. Briefly, this processing included motion correction, removal of non-brain tissue using a hybrid watershed/surface deformation procedure, automated Talairach transformation, segmentation of the subcortical white matter and deep grey matter volumetric structures, intensity normalization, tessellation of the grey matter white matter boundary, automated topology correction, and surface deformation following intensity gradients to optimally place the grey/white and grey/CSF borders at the location where the greatest shift in intensity defines the transition to the other tissue class. Once the cortical models were complete, deformable procedures were performed including surface inflation, registration to a spherical atlas based on individual cortical folding patterns to match cortical geometry across patients, parcellation of the cerebral cortex into units with respect to gyral and sulcal structure, and creation of a variety of surface-based data. This method used both intensity and continuity information from the entire MRI volume in segmentation and deformation procedures to produce representations of cortical thickness, calculated as the closest distance from the grey/white boundary to the grey/CSF boundary at each vertex on the tessellated surface. The maps were created using spatial intensity gradients across tissue classes and were therefore not simply reliant on absolute signal intensity.

All surface maps were inspected visually by a trained rater (S.R.) and scored from 1-4 based on published guidelines [24,25]. Scans with major artefacts or reconstruction errors (score >2) were excluded from further analyses. Due to the significant atrophy found on dementia with Lewy bodies scans and the impact on surface reconstruction, the cortical surfaces from dementia with Lewy bodies patients and associated controls were manually edited slice-by-slice (S.J., S.R., A.De.) and reprocessed. Cortical thickness, cortical volume, and subcortical volume measurements were next extracted from the resulting maps using the bilateral 83-region Desikan-Killiany atlas (68 cortical regions and 15 subcortical regions, namely the bilateral thalamus, caudate, putamen, pallidum, hippocampus, amygdala, nucleus accumbens, and brainstem). These metrics were all extracted because they were shown to be differentially affected in iRBD [5,26]. Given that volume scales with head size [27], volume values were normalized by dividing values by the estimated total intracranial volume. To reduce the number of input features when modelling subtypes and preserve sufficient power, the labels of each individual parcellation were fused together inside FreeSurfer to yield lobar measurements of cortical thickness for the frontal, parietal, temporal, occipital, and cingulate lobes, as done previously [16,28,29]. To control for the differences in scanner acquisitions, we next applied NeuroComBAT on the regional measurements, a batch-correcting tool widely used in multisite MRI studies that removes scanner-dependent variations while preserving the biological variance of interest, using age, sex, and disease group as covariates [30–32]. We expressed each regional measurement as a piecewise linear *z*-score normalized to the control population using age and sex as regression covariates as previously described (see Supplemental Table 2 for group descriptives) [16]. This allowed the brain measurements from each patient to reflect deviations from what was expected for age and sex, thereby ensuring that the identified progression patterns were not merely reflective of normal aging. Regions of interest were averaged between hemispheres; paired t-tests between left and right regions determined that there was no statistically significant difference between them (all p-values > 0.084). The NeuroComBAT-corrected, *z*-scored regional measurements served as the input for the analyses involving the reconstruction of transdiagnostic brain atrophy subtypes in synucleinopathies.

### Brain atrophy subtype and stage inference modelling

To reconstruct brain atrophy subtypes and stages from cross-sectional imaging data, we used the SuStaIn algorithm implemented in Python [16,33]. In contrast to conventional analyses, which would generate subtypes exclusively based on temporal progression, the SuStaIn algorithm considers both temporal and spatial information in order to define synucleinopathy groups with distinct patterns of progression (subtypes) and assigns a disease stage for each participant, thereby allowing for the identification of transdiagnostic trajectories of brain neurodegeneration. We ran SuStaIn using 25 start points and 1,000,000 Markov Chain Monte Carlo iterations. The optimal number of subtypes was determined using the cross-validation information criterion calculated through 10-fold cross-validation [16]. The SuStaIn algorithm subtyped individuals by calculating the maximum likelihood they belong to each subtype, and staged individuals by calculating their average stage weighted by the probability they belonged to each stage of each subtype. Individuals that were assigned a stage of 0 were determined to be “non-classifiable”, whereas individuals with a higher probability of belonging to a SuStaIn subtype were determined to be “classifiable”. To compare the subtype progression patterns between different neuroimaging metrics (i.e. cortical thickness versus cortical volume) and across cross-validation folds (i.e., the cross-validation similarity metric), we calculated the Bhattacharyya coefficient [34] between the position of each biomarker event in the two subtype progression patterns, averaged across biomarker events and Markov Chain Monte Carlo samples, as previously described [16]. To ensure the robustness of our subtypes and to confirm that the solution from SuStaIn was not due to having included iRBD and dementia with Lewy bodies patients, we repeated the same analyses after including a cohort of patients with Parkinson’s disease and probable RBD, processed in the same way as the other patients. We also repeated the analyses in the group of iRBD patients alone. The Bhattacharrya coefficient [34] was used to assess the similarity of these brain atrophy progression patterns compared to the initial model involving only iRBD and dementia with Lewy bodies patients. SuStaIn models were visualized using Brainpainter software [35].

### Statistical analyses

Statistical analyses were performed in R (version 4.3.2). MMSE scores were converted to MoCA scores, which involved 73 dementia with Lewy bodies and 18 iRBD patients [36]. UPDRS-III scores were converted to MDS-UPDRS-III scores in 43 iRBD patients as previously described [4]. Demographics and clinical variables were compared between patients and controls using ANOVA with post-hoc Tukey HSD testing and χ^2^ testing with post-hoc pairwise comparisons. Comparisons between subtypes used t-tests for continuous variables and χ^2^ tests for categorical variables. The progression of clinical variables with respect to stages was assessed by linear regression using age, sex, stage, subtype, and probability of subtype as covariates (i.e., clinical variable ∼ subtype + stage + age + sex + probability of subtype). Logistic regression was also used to predict phenoconversion between subtypes (i.e., phenoconversion ∼ subtype * stage + age + sex). The phenoconversion risk for each iRBD patient was calculated using this logistic regression and used for investigating the relationship with SuStaIn subtypes and stages.

## Results

### Participant demographics

Of the 1134 participants with T1-weighted imaging, 14 (1.2%) failed the FreeSurfer processing step and 157 (13.8%) did not pass surface-based quality control, leading to a final sample for analysis of 362 patients with iRBD, 82 with dementia with Lewy bodies, and 519 controls. As expected, dementia with Lewy bodies patients were older (76.8 ± 6.45 years), had lower MoCA scores (14.4 ± 5.46), and higher MDS-UPDRS-III scores (32.1 ± 18.1) compared to iRBD patients and controls. iRBD patients were younger (67.1 ± 6.95 years), with intermediate MoCA (25.7 ± 3.02) and MDS-UPDRS-III scores (6.04 ± 5.57). Controls were slightly younger than iRBD participants (65.6 ± 10.1 years) and had the highest MoCA scores (26.8 ± 2.36) and lowest MDS-UPDRS-III scores (2.28 ± 4.44). Summarized demographic and clinical information is available in Supplementary Table 2.

### SuStaIn identifies two brain atrophy subtypes

First, we used SuStaIn to identify subtypes of brain atrophy progression in the neurodegenerative spectrum linking iRBD and dementia with Lewy bodies. Since atrophy has previously been reported to be more prominent in iRBD associated with MCI and since atrophy predicts the development of dementia with Lewy bodies, we added dementia with Lewy bodies patients to our subtyping approach. These patients allowed SuStaIn to identify the brain atrophy progression patterns within the dementing spectrum linking iRBD to dementia with Lewy bodies. Using cortical thickness and subcortical volume regions of interest as input, SuStaIn identified a two-subtype model as being the best representation of brain atrophy progression in patients (Fig. 2A). This subtyping classified 247 (56%) patients with iRBD or dementia with Lewy bodies into one of the two subtypes (Fig. 2B-C), each with distinct sequences of atrophy (Fig. 2D): (i) a “cortical-first” progression subtype, found in 61% of classifiable patients, characterized by atrophy beginning in the frontal lobes followed by the temporal and parietal areas and remaining cortical areas, with the involvement of subcortical structures at later stages; and (ii) a “subcortical-first” progression subtype, found in 39% of classifiable patients, characterized by atrophy beginning in the limbic areas (primarily the amygdala and hippocampus), followed by structures of the basal ganglia and only involving cortical structures at later stages. The cortical-first subtype included 150 patients, namely 112 (75%) iRBD patients and 38 (25%) dementia with Lewy bodies patients, while the subcortical-first subtype included 97 patients, namely 67 (69%) iRBD patients and 30 (31%) dementia with Lewy bodies patients (Table 1). The remaining 197 patients with synucleinopathies (183 [93%] iRBD and 14 [7%] dementia with Lewy bodies) were categorized as stage 0/non-classifiable (i.e., assigned to very early SuStaIn stages at which point there is low confidence in the subtype assignment or displayed a different atrophy pattern compared to the rest of the sample).

**Figure 2:**
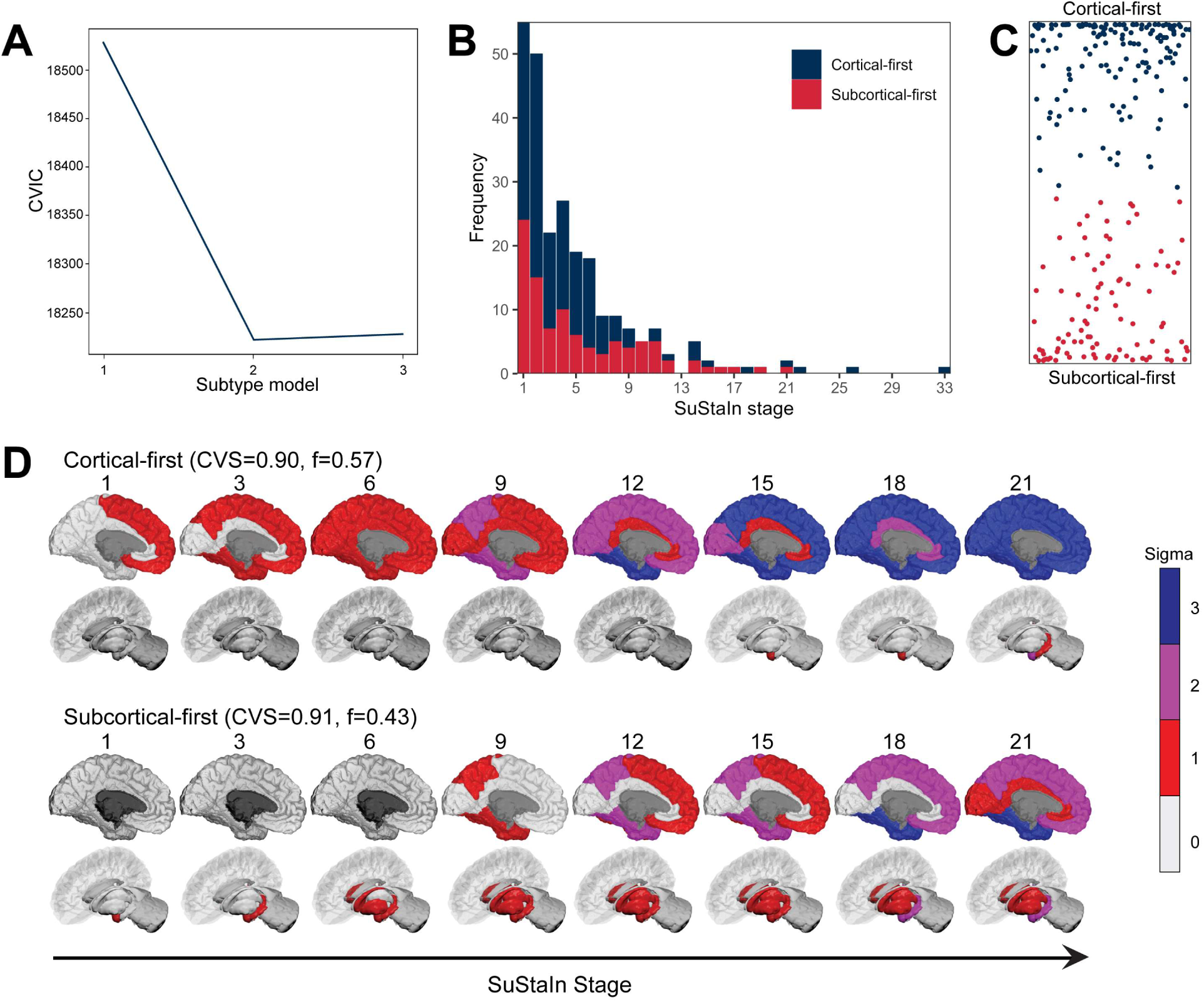
SuStaIn identified a two-subtype model as being the best representation of brain atrophy progression in patients. **(A)** CVIC across 10-fold cross-validation of left-out individuals; lower CVIC represents better model fit. **(B)** Distribution of subtypes across SuStaIn stages. **(C)** The assignability of disease subtype, operationalized as the distance from the top or bottom axis, which represents the maximum probability (100%) of that subtype. **(D)** SuStaIn identified two unique subtypes of brain atrophy progression. At each stage, the colour in each region indicates the level of severity of atrophy, with grey representing unaffected regions, red mildly affected regions (z-score of −1), magenta moderately affected regions (z-score of −2), and blue severely affected regions (z-score of −3 or more). Brainstem atrophy begins at approximately stage 6 in the subcortical-first subtype (not shown). CVIC: cross-validation information criterion; CVS: cross-validation similarity; DLB = dementia with Lewy bodies; iRBD = idiopathic/isolated REM sleep behaviour disorder; SuStaIn = Subtype and Staging Inference.

**Table 1:** Demographic and clinical variables for each brain atrophy progression subtype.

| Phenoconversion | Classifiable |  |  | Subtyped |  |  |
| --- | --- | --- | --- | --- | --- | --- |
|  | Non-classifiable | Classifiable | p-value<sup>a</sup> | Cortical-first | Subcortical-first | p-value<sup>b</sup> |
| **Demographics** |  |  |  |  |  |  |
| n (%): iRBD | 183 (92.9) | 179 (72.5) | 0.833 | 112 (74.7) | 67 (69.1) | **0.001** |
| n (%): DLB | 14 (7.1) | 68 (27.5) | **<0.001** | 38 (25.3) | 30 (30.9) | 0.332 |
| Age: All | 66.3 (7.2) | 71.0 (7.7) | **<0.001** | 70.5 (7.7) | 71.7 (7.5) | 0.249 |
| Age: iRBD | 65.8 (7.1) | 68.5 (6.5) | **<0.001** | 68 (6.3) | 69.2 (6.8) | 0.235 |
| Age: DLB | 72.7 (5.7) | 77.6 (6.3) | **<0.001** | 78 (6.7) | 77.2 (5.8) | 0.577 |
| % male | 83.8 | 83.8 | 0.960 | 82.7 | 85.6 | 0.611 |
| Stage<sup>c</sup> (SD): All | 0 (0) | 4.8 (4.7) | **<0.001** | 4.5 (4.8) | 5.3 (4.6) | 0.209 |
| Stage<sup>c</sup> (SD): iRBD | 0 (0) | 3.6 (2.9) | **<0.001** | 3.2 (2.1) | 4.3 (3.9) | **0.043** |
| Stage<sup>c</sup> (SD): DLB | 0 (0) | 7.9 (6.7) | **<0.001** | 8.2 (7.7) | 7.5 (5.4) | 0.630 |
| **Clinical variables** |  |  |  |  |  |  |
| MDS-UPDRS-III (SD): All | 6.5 (7.8) | 14.6 (16.4) | **<0.001** | 15.6 (18.2) | 13.2 (13.1) | 0.236 |
| MDS-UPDRS-III (SD): iRBD | 5.1 (4.7) | 7.1 (6.3) | **<0.001** | 7.1 (6.5) | 7.0 (5.9) | 0.932 |
| MDS-UPDRS-III (SD): DLB | 25.2 (14.2) | 33.5 (18.6) | 0.073 | 38.7 (19.9) | 26.9 (14.5) | **0.006** |
| MoCA (SD): All | 25.6 (3.5) | 22.5 (6.3) | **<0.001** | 22.5 (6.7) | 22.5 (5.8) | 0.955 |
| MoCA (SD): iRBD | 26.1 (2.8) | 25.4 (3.2) | **0.040** | 25.4 (3.3) | 25.5 (2.9) | 0.855 |
| MoCA (SD): DLB | 17.7 (5.1) | 13.8 (5.4) | **0.048** | 12.9 (5.9) | 15.0 (4.4) | 0.112 |
| % MCI: iRBD<sup>d</sup> | 35.6 | 46.3 | **0.041** | 50.0% | 48.5% | 0.649 |

Statistical differences were calculated using unpaired t-tests for continuous variables and chi-squared test for categorical variables.
<sup>a</sup>Non-classifiable group versus classifiable group.
<sup>b</sup>Cortical-first subtype versus subcortical-first subtype.
<sup>c</sup>Stage refers to SuStaIn stage.
<sup>d</sup>MCI as defined by $\leq 25/30$ on MoCA; all DLB patients met criteria for dementia.
DLB = dementia with Lewy bodies; iRBD = idiopathic/isolated REM sleep behaviour disorder; MoCA = Montreal Cognitive Assessment; MCI = mild cognitive impairment; MDS-UPDRS-III = Movement Disorders Society – Unified Parkinson's Disease Rating Scale, Part III; MoCA = Montreal Cognitive Assessment; PD-pRBD = Parkinson's disease with possible REM sleep behaviour disorder; SD = standard deviation; SuStaIn = Subtype and Staging Inference.

The average similarity between cross-validation folds was >90% for each subtype, indicating high reliability of subtype progression patterns with 10-fold cross-validation. Moreover, the identification of two distinct subtypes was recapitulated when using cortical volume (as a measure of cortical atrophy instead of cortical thickness) with subcortical volume as input features, with 86% similarity when comparing the subtypes’ progression patterns (Supplementary Figure 1). To ensure the robustness of our subtyping, we performed secondary analyses by including Parkinson’s disease patients with probable RBD as inputs into the SuStaIn modeling, as well as by using iRBD patients alone (Supplementary Figure 2 and Supplementary Table 3). In both cases, the two subtypes identified in the primary SuStaIn model were recapitulated with similar patterns of progression, although as expected, there was increased uncertainty at higher stages in the iRBD-only model, particularly in the cortical-first subtype. As with the main SuStaIn model, the inclusion of Parkinson’s disease patients with probable RBD resulted in a similar distribution of classifiable patients (304 classifiable versus 250 non-classifiable patients), with 50% of Parkinson’s disease patients determined to be non-classifiable. In both cases, the Bhattacharrya coefficient indicated a similarity between 81% and 90% with the original model that included iRBD and dementia with Lewy bodies patients. Taken together, this indicates that the primary driver of subtyping and staging reflects the progression of cortical and subcortical atrophy.

Inspecting the subtypes based on the progression of atrophy in each brain region revealed that compared to normative data from control scans, iRBD patients from the subcortical-first subtype had rapid subcortical volume loss in the early stages, with relative stability of most cortical structures but progressive atrophy of the hippocampus, putamen, and cortical structures at later stages (Figure 3). This pattern was generally reversed in cortical-first patients, where atrophy of cortical structures occurred in the earlier stages followed by relative stability in the cingulate, occipital, and parietal structures, with progressive atrophy in the frontal, insular, and temporal cortical areas and subcortical structures (Figure 3).

**Figure 3:**
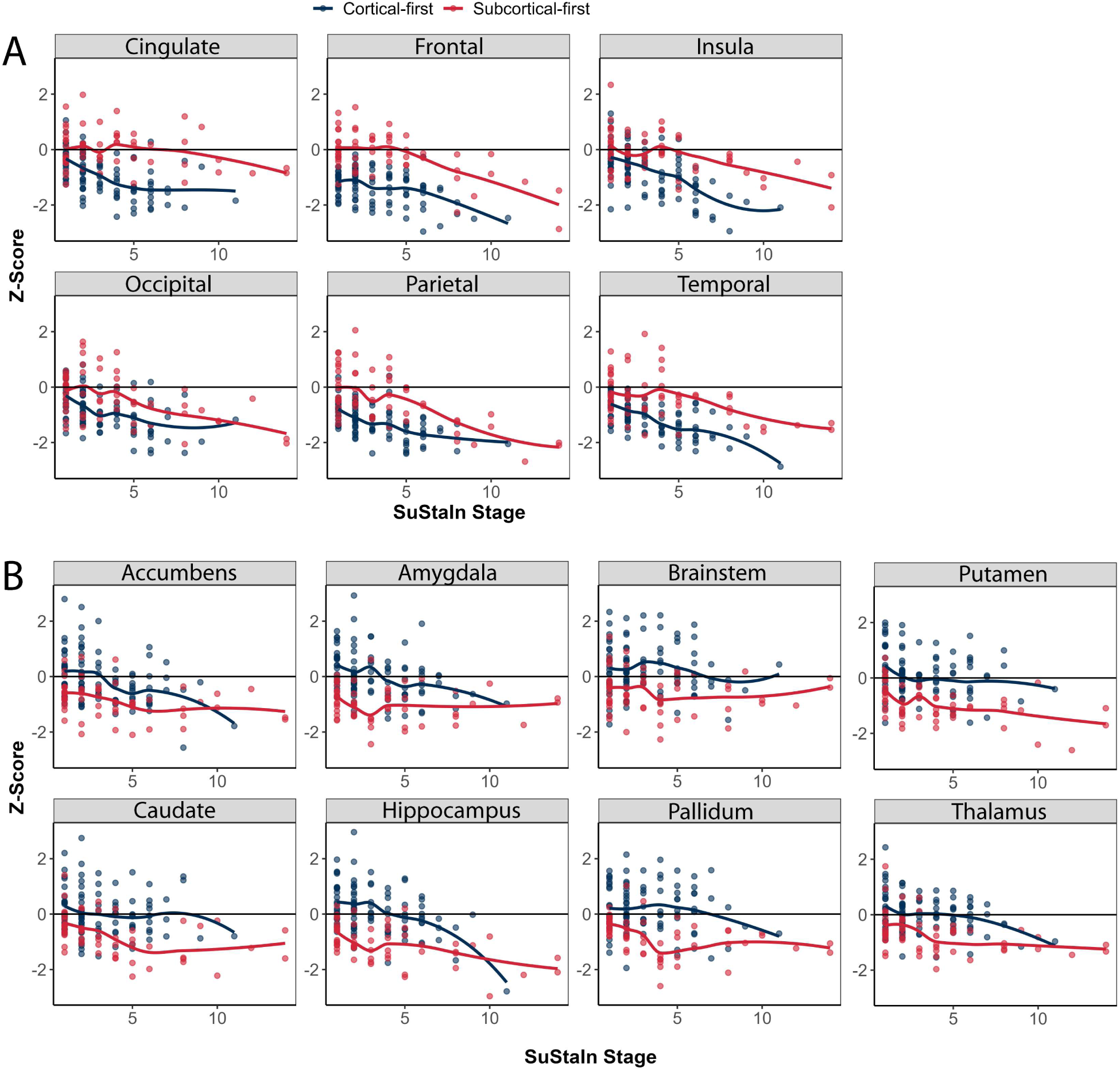
Progression of cortical and subcortical atrophy by subtype and stage in iRBD. The progression of atrophy in cortical regions (**A**) and subcortical regions (**B**) used in the SuStaIn modelling in classifiable iRBD patients. iRBD = idiopathic/isolated REM sleep behaviour disorder.

### Atrophy subtypes are related to increased clinical burden

Next, we investigated whether demographics and clinical variables differed between classifiable and stage 0/non-classifiable patients and between the identified subtypes. The baseline demographics and clinical variables of the classifiable and stage 0/non-classifiable groups are shown in Table 1. The classifiable group (which includes patients identified as either cortical-first or subcortical-first subtypes) had more dementia with Lewy bodies patients (27.5% *vs*. 7.1%, p < 0.001), were overall older (71.0 ± 7.7, *vs.* 66.3 ± 7.2 p < 0.001), and had worse MoCA (22.5 ± 6.3 *vs.* 25.6 ± 3.5, p < 0.001), and MDS-UPDRS-III (14.6 ± 16.4 *vs*. 6.5 ± 7.8, p < 0.001) scores than stage 0/non-classifiable patients. Worse clinical scores in classifiable patients were also observed when comparing within iRBD and dementia with Lewy bodies groups.

There were no significant differences in sex proportion, age, and MoCA scores when comparing patients classified in the cortical-first versus the subcortical-first atrophy progression subtypes (Table 1). However, there were significantly more iRBD patients classified as cortical-first than subcortical-first (74.7% *vs.* 69.1%, p = 0.001), whereas the distribution in dementia with Lewy bodies patients was not significantly different (25.3% *vs.* 30.9%, p = 0.332). In addition, although the MDS-UPDRS-III scores were not different between subtypes in the total sample of patients, those with dementia with Lewy bodies classified in the cortical-first group had higher MDS-UPDRS-III scores than those classified in the subcortical-first subtype (38.7 ± 19.9 versus 26.9 ± 14.5, p = 0.006). In other words, our modelling identified brain atrophy subtypes related to higher cognitive and motor disease burden.

### Brain atrophy severity relates to cognitive and motor progression

We next sought to determine whether cognitive and motor functions varied as a function of brain subtype severity. Figure 4 and Table 2 show the relationships between clinical variables and SuStaIn subtype and stage while accounting for age, sex, and probability of subtype. Higher SuStaIn stages, namely more advanced brain disease progression, were associated with higher MDS-UPDRS-III scores (increased by 0.69 points per increase in stage, p = 0.003) and lower MoCA scores (decreased by 0.35 points per increase in stage, p < 0.001). Moreover, the cortical-first subtype was associated with a greater rate of increase in MDS-UPDRS-III scores over time relative to the subcortical-first subtype (increased by 5.03 points, p = 0.011). This indicates that as brain atrophy progresses, cognitive and motor impairments become more pronounced, with the cortical-first subtype exhibiting a greater motor disease burden compared to the subcortical-first subtype.

**Figure 4:**
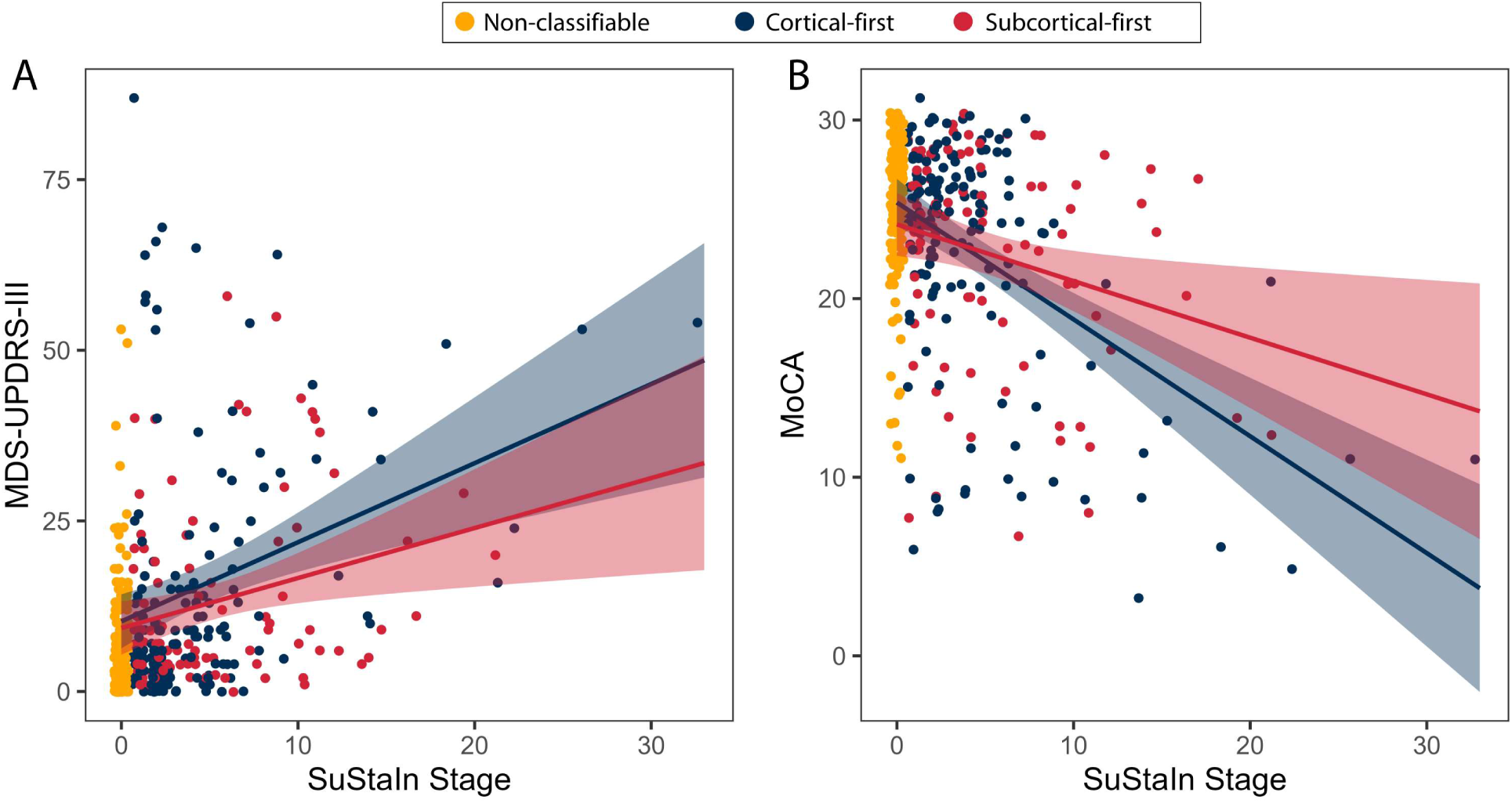
Progression of clinical variables by SuStaIn stage. Higher SuStaIn stages was associated with worse clinical scores on MDS-UPDRS-III (**A**) and MoCA (**B**) in iRBD and DLB patients. DLB = dementia with Lewy bodies; iRBD = idiopathic/isolated REM sleep behaviour disorder; MoCA = Montreal Cognitive Assessment; MDS-UPDRS-III = Movement Disorders Society – Unified Parkinson’s Disease Rating Scale, Part III; SD = standard deviation.

**Table 2:** Comparison of clinical variables and time to phenoconversion by SuStaIn subtype and stage.

|  | SuStaIn subtype |  | SuStaIn stage |  |
| --- | --- | --- | --- | --- |
|  | Beta estimate | p-value | Beta estimate | p-value |
| **Clinical variables** |  |  |  |  |
| MDS-UPDRS-III | -5.03 | **0.011** | 0.69 | **0.003** |
| MoCA | 1.06 | 0.144 | -0.35 | **<0.001** |

Linear regression model of variable ~ stage + subtype + age + sex + probability of subtype.
iRBD = idiopathic/isolated REM sleep behaviour disorder; MoCA = Montreal Cognitive Assessment; MDS-UPDRS-III = Movement Disorders Society – Unified Parkinson’s Disease Rating Scale, Part III; SD = standard deviation; SuStaIn = Subtype and Staging Inference.

### Atrophy subtypes relate differently to phenoconversion in iRBD

We investigated whether the brain atrophy subtypes are associated with phenoconversion risk and clinical subtypes in iRBD. Among all iRBD patients, 84 (23%) phenoconverted to a defined synucleinopathy, with 28 (33%) having developed dementia with Lewy bodies, 52 (62%) Parkinson’s disease, and 4 (5%) multiple system atrophy. Although there were no differences in the unadjusted number of phenoconverted patients between classifiable and non-classifiable groups or between subtypes (Supplementary Table 4), we further analysed the risk of phenoconversion in iRBD with respect to classifiability using a logistic regression model involving age, sex, and interaction between classifiability and probability of classifiability (Supplementary Table 5). This revealed that classifiable subjects (either cortical-first or subcortical-first) had a higher predicted probability of phenoconverting as compared to non-classifiable subjects (log-odds = 3.38, p = 0.040). This effect was modulated by the interaction between classifiability and probability of subtype (log-odds = −4.32, p = 0.034), since most non-classifiable subjects had lower probability of subtyping as compared to classifiable subjects.

To analyse the risk of phenoconversion in iRBD with respect to a specific SuStaIn subtype, we employed a logistic regression model using age, sex, and interaction between subtype and stage (Supplementary Table 6). This revealed a significant effect of subtype on phenoconversion, with iRBD patients in the subcortical-first subtype being more likely to phenoconvert compared to those classified in the cortical-first subtype (log-odds = 1.45, p = 0.022). Importantly, the interaction term between SuStaIn subtype and stage was also significant (log-odds = −0.31, p = 0.049), indicating that the effect of severity of atrophy on the likelihood of phenoconverting differed between the iRBD patients classified in each subtype. Specifically, compared to iRBD patients classified in the cortical-first subtype, iRBD patients classified in the subcortical-first subtype had a higher likelihood of receiving a diagnosis of an overt synucleinopathy in the earliest severity stages of brain disease progression.

Subtype-specific logistic regression models indicated that an earlier SuStaIn stage in iRBD in the subcortical-first subtype related to a higher likelihood of phenoconverting to an overt disease (log-odds = −0.232, p = 0.054) independently from age (p = 0.75) and sex (p = 0.88) (Supplementary Table 6). Conversely, in the iRBD patients classified as cortical-first, older age was significantly associated with a higher likelihood of phenoconversion (log-odds = 0.082, p = 0.049) but not SuStaIn stage (p = 0.62) or sex (p = 0.52) (Supplementary Table 7). In other words, more prominent brain atrophy in subcortical structures early during the disease course was associated with a stronger likelihood of phenoconverting at earlier stages (Figure 5A), resulting in a clinical diagnosis of a defined neurodegenerative disease sooner. On the other hand, more prominent brain atrophy in cortical structures early during the disease course was associated with phenoconversion over the long term (Figure 5A), with these patients receiving a clinical diagnosis at much later stages of brain disease progression.

**Figure 5:**
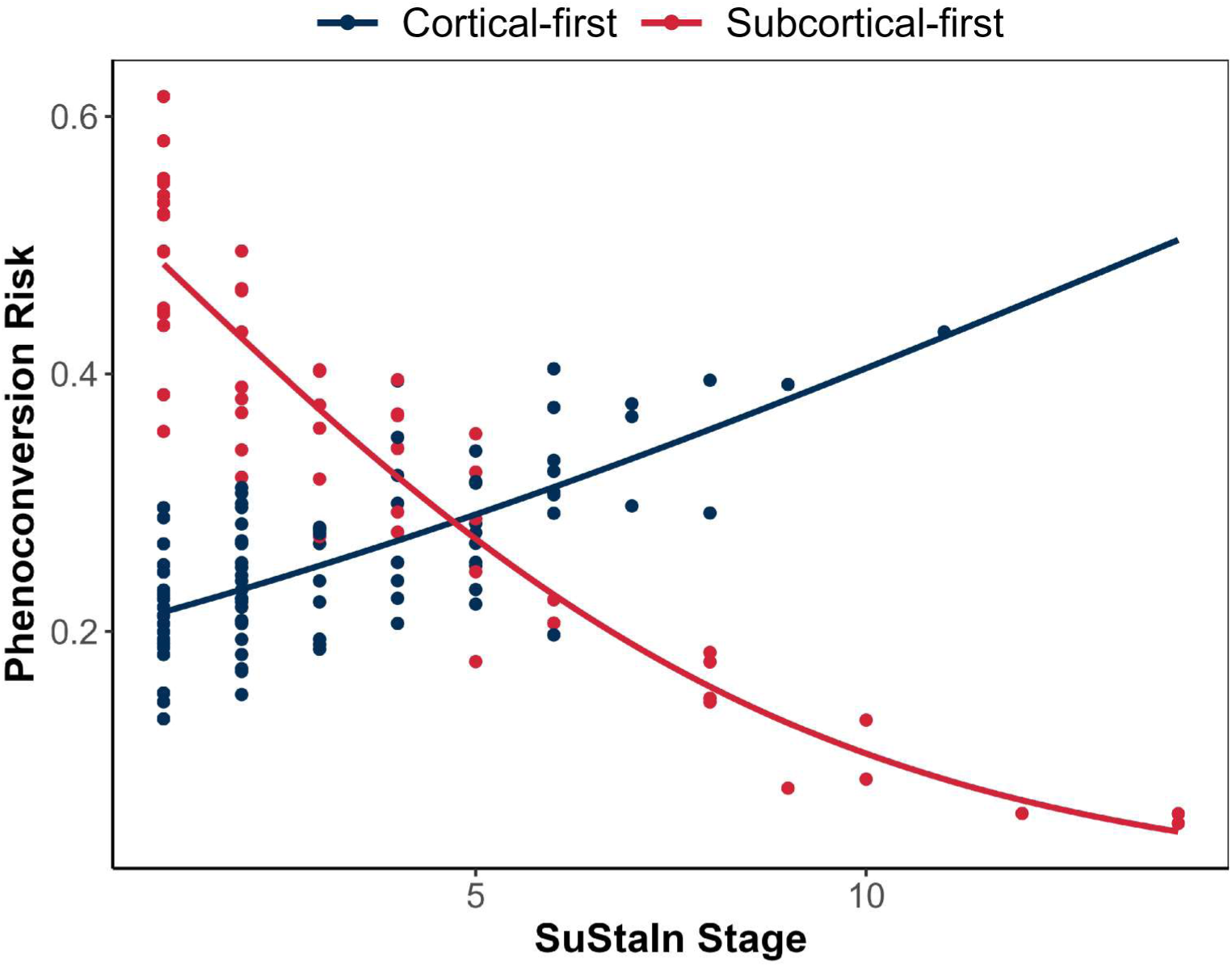
Phenoconversion risk (calculated from the logistic regression predicting phenoconversion) differs in iRBD patients based on SuStaIn subtype and stage. iRBD patients classified in the subcortical-first subtype had a stronger likelihood of phenoconverting to overt disease at earlier stages (and less in later stages as diagnosis was more likely to occur earlier), whereas iRBD patients classified in the cortical-first subtype had a stronger likelihood of phenoconversion over the long term. iRBD = idiopathic/isolated REM sleep behaviour disorder; SuStaIn = Subtype and Staging Inference.

We then investigated whether the subtypes could serve as predictors for the development of a parkinsonism or dementia phenotype while still in the preclinical stage of iRBD, as predicting differential pathways from still healthy iRBD individuals is essential for developing a prognostic subtyping approach. Logistic regression revealed that the development of a parkinsonism phenotype in iRBD patients was associated with the subcortical-first subtype compared to those who did not yet phenoconvert (log-odds = 0.63, p = 0.014), but not with SuStaIn stage (p = 0.21), age (p = 0.61), or sex (p = 0.97) (Supplementary Table 8). In contrast, the development of dementia in iRBD patients was borderline-significantly associated with SuStaIn stage (log-odds = 0.37, p = 0.054), independently of subtype (p = 0.77), age (p = 0.37), or sex (p = 0.90) (Supplementary Table 8). These findings indicate that classification into a subcortical-first subtype is associated with a higher likelihood of developing a parkinsonism-first phenotype. In addition, the progression of brain atrophy, regardless of subtype, is related to an increased risk of developing dementia.

## Discussion

In this study, we used a data-driven approach to identify two distinct patterns of brain atrophy progression, summarizing the spectrum linking iRBD to overt disease. The first is a cortical-first atrophy progression subtype, where atrophy initially spreads throughout cortical areas before manifesting in subcortical structures later. The second is a subcortical-first atrophy progression subtype, where atrophy begins in the amygdala and basal ganglia before spreading to the cortical areas. Patients classified in the subtypes had an increased clinical burden compared to patients not subtyped by our modelling. Clinical scores of disease severity worsened with increasing stages of atrophy, and phenoconversion trajectories differed based on the subtype. Specifically, iRBD patients with a subcortical-first atrophy subtype were more likely to phenoconvert at earlier SuStaIn stages, while those with a cortical-first pattern of atrophy were more likely to phenoconvert at later stages. In addition, iRBD patients with a subcortical-first atrophy subtype were more likely to phenoconvert to a parkinsonism-first phenotype, while both subtypes were likely to phenoconvert to a dementia-first phenotype. Our results provide insights into the progression of brain atrophy in prodromal synucleinopathy as it develops towards manifest disease, which may have potential utility for prognostication and patient stratification.

Previous studies have found cortical and subcortical atrophy in iRBD patients, which have been shown to correlate with motor and cognitive dysfunction, as well as predict phenoconversion to dementia [5,7–9,15,37]. The atrophy in iRBD, as in several neurodegenerative diseases [13,38,39], has been shown to be constrained by both the brain’s structural connectivity pattern and the local patterns of gene expression [5], targeting preferentially regions overexpressing genes involved in energy production and protein degradation [6]. Distinct patterns of cortical and subcortical atrophy have also been described in patients with mild cognitive impairment who later developed dementia with Lewy bodies [40]. Patients with dementia with Lewy bodies similarly show unique patterns of atrophy when compared with patients with Alzheimer’s disease and healthy controls [41,42], with a hippocampal-sparing pattern of regional atrophy observed in dementia with Lewy bodies, which may be influenced by mixed co-pathology [43]. The distinct involvement of subcortical structures at earlier and later disease stages depending on machine learning-derived subtypes has also been described individuals with manifest Parkinson’s disease [44]. The broad areas and patterns of atrophy in prodromal synucleinopathy and overt disease found in the present study are in line with these results. Here, using a large cross-sectional sample size of brain MRI scans in iRBD and dementia with Lewy bodies and machine learning, we were able to account for the variability in disease stage across individuals and reconstruct the progression of atrophy even at very early stages of disease. Our results not only support the finding that atrophy is diffuse in the late stages of synucleinopathy, but also suggest that the origin and pathway towards this state follows distinct patterns. These different patterns of atrophic spread, based solely on the data-driven analysis of quantitative atrophy derived from brain MRI scans, could have relevance for prognosis or more precisely select patients for disease-modifying trials. For this to be the case, future studies will need to derive signature patterns for each of these subtypes and develop algorithms that will allow classifying brain MRI scans from patients into the likeliest subtype.

The identified subtypes were significantly associated with clinical features and progression trajectories. Indeed, we observed that higher subtype stages, namely more advanced brain disease progression, were associated with worse clinical scores. Furthermore, the cortical-first subtype was associated with higher rate of increase in MDS-UPDRS-III scores over time relative to the subcortical-first subtype. This agrees with our observation that cortical-first patients phenoconvert at later stages, having time to accumulate more brain changes as they transition towards overt disease. This is in line with both dementia with Lewy bodies and Parkinson’s disease phenoconverters having similarly elevated MDS-UPDRS-III in the iRBD stage, and that the motor interval being, if anything, longer in dementia with Lewy bodies phenoconverters than Parkinson’s disease phenoconverters [45]. Importantly, the MDS-UPDRS-III and MoCA are broad metrics of motor and cognitive function, respectively, which do not fully capture the breadth or depth of dysfunction in iRBD [45,46]. Future work can examine if different subtype progression patterns are associated with more specific patterns of clinical dysfunction.

Regression analyses indicated that classifiable subjects with iRBD had higher risk of phenoconversion than non-classifiable subjects. Furthermore, the subcortical-first brain atrophy progression subtype in iRBD was associated with a stronger likelihood of developing an overt synucleinopathy at earlier stages, whereas the cortical-first brain atrophy progression subtype was associated with phenoconversion over the longer term. We propose that as subcortical structures are affected initially, the hallmark clinical features of parkinsonism become manifest, leading to a diagnosis at earlier subcortical stages. In line with this, phenoconversion occurs at later stages in the cortical-first subtype, since it takes longer for the subcortical structures to be affected and motor signs and symptoms of disease to become manifest. This is supported by the fact that: i) the subcortical-first subtype was associated with conversion to the parkinsonism phenotype, particularly at earlier stages; ii) the cortical-first subtype was associated with a greater rate of increase in MDS-UPDRS-III over time, which implicitly suggests that subcortical-first phenoconverters are at higher scores initially with less room to progress; and iii) conversion to the dementia phenotype occurred at later SuStaIn stages irrespective of subtype. In other words, whereas both subtypes are associated with dementia with increasing progression, the subcortical-first phenotype is more strongly associated with the development of a parkinsonism subtype in iRBD (Figure 6). This may indicate that the cortical-first subtype is more closely related to what is classically known as dementia with Lewy bodies (i.e., initial cortical involvement followed by subcortical involvement, with a long-term risk of dementia), whereas the subcortical-first subtype is more closely related to Parkinson’s disease with dementia (i.e., initial subcortical involvement followed by cortical involvement, with earlier phenoconversion to Parkinson’s disease and an increased long-term risk of dementia).

**Figure 6:**
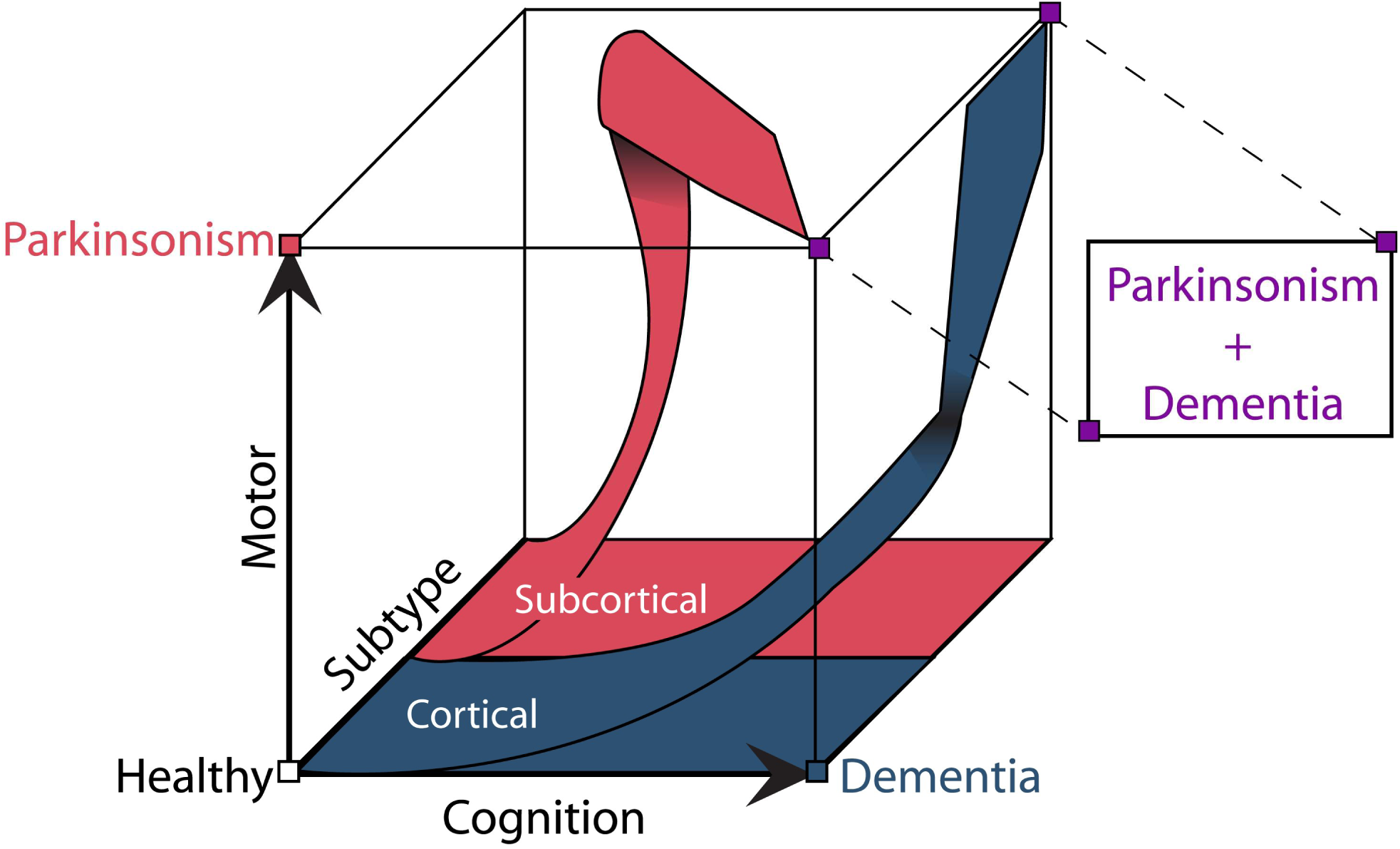
Hypothetical schematic representing the pathways of evolution of brain atrophy progression in iRBD, as simulated by SuStaIn. In this model, the subcortical-first subtype is associated with earlier phenoconversion to parkinsonism, possibly due to initial involvement of the basal ganglia structures, with the development of dementia only at later stages of disease. By contrast, the cortical-first subtype is associated with phenoconversion only at later stages of disease progression. This model suggests that the cortical-first subtype is more closely related to what is classically known as DLB (i.e., initial cortical involvement followed by subcortical involvement, with a long-term risk of dementia), whereas the subcortical-first subtype is more closely related to PD with dementia (i.e., initial subcortical involvement followed by cortical involvement, with earlier phenoconversion to PD and an increased long-term risk of dementia). DLB = dementia with Lewy bodies; iRBD = idiopathic/isolated REM sleep behaviour disorder; PD = Parkinson’s disease.

Several hypotheses may explain the pathophysiological patterns of each subtype. First, the patterns of atrophy may be reflective of ongoing neurodegeneration in different regions of the brain in iRBD: the cortical-first subtype begins with neurodegeneration of the cortex while the subcortical-first subtype begins with atrophy in the limbic and basal ganglia structures. It is important to keep in mind that our model was built on atrophy and not actual pathology, and that although preliminary evidence has shown that atrophy in synucleinopathies can be recreated *in silico* as a spread of alpha-synuclein misfolded proteins [5], several other proteins and co-pathologies may also be at play in iRBD-associated neurodegeneration. For example, brain neurodegeneration in dementia with Lewy bodies patients is associated with amyloid beta and tau deposition at baseline [47,48] and lower CSF levels of amyloid beta 42 have been found in iRBD compared to controls [49]. Moreover, 25% of iRBD patients are found to be amyloid beta-positive [50]. Future studies should investigate whether the cortical- and subcortical-first atrophy subtypes of iRBD differ on imaging and blood- and CSF-based markers of Alzheimer’s disease co-pathologies. Another possibility is that the cortical-first subtype represents a resilient subtype compared to the subcortical-first subtype, where pathology also spreads through the subcortical structures but did not manifest as observable and quantifiable atrophy, unlike cortical areas. From this angle, both subtypes would have the same initial starting point, and patients within the subcortical-first subtype would represent increased vulnerability of the basal ganglia structures in showing neurodegeneration and displaying atrophy. Otherwise, despite iRBD patients being classified in this study as cortical- or subcortical-first, previous models have demonstrated that the iRBD phenotype belongs to a body-first propagation of pathology compared to a brain-first (i.e., pathology spreading from the gut to the brain and not from the brain to the gut) [51]. Therefore, it could be that the impact of pathology differs between subtypes, with the cortical-first subtype impacting more strongly upon several brainstem nuclei and neurotransmitter systems whose upstream changes yield observable morphological changes. Finally, it may be that deviations in morphological measurements compared to what was expected for age and sex are reflective of long-term genetic, lifestyle and environmental factors, which render the brain differently vulnerable to synucleinopathies once the pathological process hits.

Approximately half of the iRBD patients were not classifiable into a disease subtype. The stage 0/non-classifiable patients were significantly younger, had better MDS-UPDRS-III and MoCA scores, and less overall brain atrophy. This was expected, as previous studies using computational neuroimaging in iRBD demonstrated that cognitive impairments account for a large variance of the morphological changes associated with iRBD [8], being significantly more prominent in the presence of mild cognitive impairment [7], which affects 30% to 50% of iRBD patients [10,52]. A smaller number of dementia with Lewy bodies patients (17%) were non-classifiable, similar in proportion to a recent SuStaIn study in patients with progressive supranuclear palsy [29]. Dementia with Lewy bodies patients that were non-classifiable were also younger relative to classifiable dementia with Lewy bodies patients and had better MoCA scores, although all met criteria for dementia. It is possible that non-classifiable iRBD and dementia with Lewy bodies patients reflect phenotypes with less overall disease burden or perhaps different patterns of brain atrophy progression. For example, it is known that *de novo* dementia with Lewy bodies patients have much higher frequencies of significant Alzheimer’s disease co-pathology as compared to *de novo* Parkinson’s disease patients, implying that Alzheimer’s disease co-pathology during the prodromal phase is a strong determinant of dementia with Lewy bodies phenoconversion and the development of dementia in synucleinopathies in general [53]. Information about Alzheimer’s disease co-pathology was not available for this study; however, we speculate that the unclassifiable dementia with Lewy bodies and iRBD patients could reflect a “pure” alpha-synuclein phenotype with limited co-pathology, with consequently a lesser degree of atrophy. In keeping with this possibility, adding Parkinson’s disease patients with probable RBD – who had less overall atrophy and presumably less Alzheimer’s disease co-pathology – to the SuStaIn model did not substantively change the subtyping patterns, nor did it result in a significant number of previously non-classifiable patients becoming classifiable or create a novel alternative subtype. Moreover, 50% of the Parkinson’s disease patients were not classifiable, in keeping with the fact that atrophy was the primary driver of subtyping and staging. Once a computational framework becomes available for obtaining a probability of subtyping from individual brain MRI scans in iRBD, future studies should investigate more thoroughly the clinical features and biological underpinnings of classified and unclassified patients.

Some limitations in this study should be discussed. First, the modelling based on the SuStaIn algorithm recreated spatiotemporal brain atrophy progression patterns from cross-sectional MRI scans. Although powerful for leveraging large datasets of brain disease scans, future initiatives should aim at investigating the differential pathways of brain disease progression from longitudinal MRI scans in iRBD patients. Second, even though this multicentric study involved the largest MRI sample of patients of polysomnography-proven iRBD, the number of patients remains limited, which increases the uncertainty of staging. Moreover, large regions of interest were used to better balance the spatial and temporal dimensions, which may have hidden the presence of atrophy in smaller areas. This is even more important for the brainstem, where specific nuclei have been reported to be impacted by neurodegeneration and pathology in iRBD [54–56]. Another limitation of the study is the combination of patients from different centres including distinct imaging and acquisition protocols. However, we performed our analyses on imaging data that were harmonized for the effect of imaging site using NeuroComBAT. All dementia with Lewy bodies diagnoses were made clinically and not confirmed at post-mortem; thus, some degree of misdiagnosis cannot be excluded, for example, with Alzheimer’s disease. Furthermore, dementia with Lewy bodies patients were also not polysomnography-proven to have RBD, although RBD is highly prevalent in dementia with Lewy bodies and is a core clinical feature in the diagnosis [22]. In order to harmonize clinical data, MMSE scores were converted to estimated MoCA scores in a subset of participants, which may limit the interpretation of cognitive function since the MMSE is less sensitive to mild cognitive impairment [36]. However, the majority of such conversions involved those with dementia with Lewy bodies, who meet criteria for dementia by definition. Finally, due to limitations on available MRI studies, we were not able to verify subtyping patterns using an independent replication sample set; however, to our knowledge, our primary analysis has used by far the largest sample size of prodromal synucleinopathy MRIs assembled to date.

## Conclusions

In conclusion, we demonstrate data-driven evidence for the existence of two atrophy subtypes in iRBD. The accurate identification and staging of patients with iRBD may have important implications for tracking disease progression.

## Data Availability

The datasets used and/or analysed during the current study are available from the corresponding author on reasonable request. Source code for the pySuStaIn algorithm is available at http://github.com/ucl-pond/.

## List of abbreviations

CCNA: Canadian Consortium on Neurodegeneration in Aging
iRBD: isolated/idiopathic REM sleep behaviour disorder
MDS-UPDRS-III: Movement Disorders Society – Unified Parkinson’s Disease Rating Scale part III
MMSE: Mini-Mental State Exam
MoCA: Montreal Cognitive Assessment
PPMI: Parkinson’s Progression Markers Initiative
SuStaIn: Subtype and Stage Inference

## Declarations

### Consent for publication

Not applicable

## Acknowledgements

S.J. was supported by an Edmond J. Safra Fellowship in Movement Disorders from the Michael J. Fox Foundation. P.C.D. is funded by the Medical Research Council [grant number MR/W000229/1]. J.F.G. reports grants from the Fonds de recherche du Québec – Santé, the Canadian Institutes of Health Research, the W. Garfield Weston Foundation, the Michael J. Fox Foundation for Parkinson’s Research, and the National Institutes of Health. R.B.P. reports grants from the Canadian Institute of Health Research, the Michael J. Fox Foundation, the Webster Foundation, Roche, and the National Institute of Health as well as personal fees from Takeda, Biogen, Abbvie, Curasen, Lilly, Novartis, Eisai, Paladin, Merck, Korro, Vaxxinity, Bristol Myers Squibb, and the International Parkinson and Movement Disorders Society, outside the submitted work. H.C. is supported by a Chair in Cognitive Neurology and Innovation from the Baycrest Academy for Research and Education, Baycrest Health Sciences, Toronto. He has been supported by a Foundation Grant from the Canadian Institutes for Health Research, along with operating funds from the Weston Foundation, Weston Brain Institute, and the National Institute of Health. He is Scientific Director (unpaid) for the Canadian Consortium on Neurodegeneration in Aging. S.L. is supported by a Leadership Fellowship from the National Health and Medical Research Council (#1195830). E.M. reports funding from the National Health and Medical Research Council (#2008565) and US department of Defense Congressionally Directed Medical Research Program Early Investigator Grant (PD220061). K.A.E.M. reports funding from Parkinson Canada, Parkinson’s Movement Disorder Foundation, University of Waterloo International Research Partnership grant, and Natural Science and Engineering Research Council of Canada. L.C. is the recipient of the Bierzonski Burczyk Foundation Postgraduate Research Scholarship. B.O. received a research grant from G.E. Healthcare. S.R. reports grants from Alzheimer Society Canada, Brain Canada, and Parkinson Canada.

Data used in the preparation of this article were partly obtained from the Canadian Consortium on Neurodegeneration in Aging and the Parkinson’s Progression Markers Initiative databases. The Canadian Consortium on Neurodegeneration in Aging and the Comprehensive Assessment of Neurodegeneration and Dementia (COMPASS-ND) study were supported by the Canadian Institutes for Health Research (#CNA-137794, #CNA-163902, #BDO-148341) along with partner support from a set of partners including not for profit organizations: Brain Canada, Alzheimer Society of Canada, Women’s Brain Health Initiative, Picov Family Foundation, New Brunswick Health Research Foundation, Saskatchewan Health Research Foundation, and Ontario Brain Institute. R.C. is team Lead of Team 8 (Lewy body disease) of the Canadian Consortium for Neurodegeneration of Aging.

The Parkinson’s Progression Markers Initiative (PPMI)—a public-private partnership—is funded by the Michael J. Fox Foundation for Parkinson’s Research and funding partners, including 4D Pharma, AbbVie Inc., AcureX Therapeutics, Allergan, Amathus Therapeutics, Aligning Science Across Parkinson’s (ASAP), Avid Radiopharmaceuticals, Bial Biotech, Biogen, BioLegend, Bristol Myers Squibb, Calico Life Sciences LLC, Celgene Corporation, DaCapo Brainscience, Denali Therapeutics, The Edmond J. Safra Foundation, Eli Lilly and Company, GE Healthcare, GlaxoSmithKline, Golub Capital, Handl Therapeutics, Insitro, Janssen Pharmaceuticals, Lundbeck, Merck & Co., Inc., Meso Scale Diagnostics, LLC, Neurocrine Biosciences, Pfizer Inc., Piramal Imaging, Prevail Therapeutics, F. Hoffman-La Roche Ltd and its affiliated company Genentech Inc., Sanofi Genzyme, Servier, Takeda Pharmaceutical Company, Teva Neuroscience, Inc., UCB, Vanqua Bio, Verily Life Sciences, Voyager Therapeutics, Inc., and Yumanity Therapeutics, Inc. For up-to-date information on the study, visit www.ppmi-info.org.

## Appendix 1

List of the contributors involved in the ICEBERG Study Group:

Steering committee: Marie Vidailhet, MD, PhD, (Pitié-Salpêtrière Hospital, Paris, principal investigator of ICEBERG), Jean-Christophe Corvol, MD, PhD (Pitié-Salpêtrière Hospital, Paris, scientific lead), Isabelle Arnulf, MD, PhD (Pitié-Salpêtrière Hospital, Paris, member of the steering committee), Stéphane Lehericy, MD, PhD (Pitié-Salpêtrière Hospital, Paris, member of the steering committee);

Clinical data: Marie Vidailhet, MD, PhD, (Pitié-Salpêtrière Hospital, Paris, coordination), Graziella Mangone, MD, PhD (Pitié-Salpêtrière Hospital, Paris, co-coordination), Jean-Christophe Corvol, MD, PhD (Pitié-Salpêtrière Hospital, Paris), Isabelle Arnulf, MD, PhD (Pitié-Salpêtrière Hospital, Paris), Smaranda Leu MD (Pitié-Salpêtrière Hospital, Paris), Sara Sambin, MD (Pitié-Salpêtrière Hospital, Paris), Jonas Ihle, MD (Pitié-Salpêtrière Hospital, Paris), Caroline Weill, MD, (Pitié-Salpêtrière Hospital, Paris), Poornima MENON MD, (Pitié-Salpêtrière Hospital, Paris), David Grabli, MD, PhD (Pitié-Salpêtrière Hospital, Paris); Florence Cormier-Dequaire, MD (Pitié-Salpêtrière Hospital, Paris); Louise Laure Mariani, MD, PhD (Pitié-Salpêtrière Hospital, Paris), Emmanuel Roze, MD, PhD, (Pitié-Salpêtrière Hospital, Paris), Cécile Delorme, MD (Pitié-Salpêtrière Hospital, Paris), Elodie Hainque MD, PhD, (Pitié-Salpêtrière Hospital, Paris), Aurelie Méneret (MD, PhD, (Pitié-Salpêtrière Hospital, Paris), Bertrand Degos, MD, PhD (Avicenne Hospital, Bobigny);

Neuropsychological data: Richard Levy, MD (Pitié-Salpêtrière Hospital, Paris, coordination), Fanny Pineau, MS (Pitié-Salpêtrière Hospital, Paris, neuropsychologist), Julie Socha, MS (Pitié-Salpêtrière Hospital, Paris, neuropsychologist), Eve Benchetrit, MS (La Timone Hospital, Marseille, neuropsychologist), Virginie Czernecki, MS (Pitié-Salpêtrière Hospital, Paris, neuropsychologist), Marie-Alexandrine Glachant, MS (Pitié-Salpêtrière Hospital, Paris, neuropsychologist);

Eye movement: Sophie Rivaud-Pechoux, PhD (ICM, Paris, coordination); Elodie Hainque, MD, PhD (Pitié-Salpêtrière Hospital, Paris);

Sleep assessment: Isabelle Arnulf, MD, PhD (Pitié-Salpêtrière Hospital, Paris, coordination), Smaranda Leu Semenescu, MD (Pitié-Salpêtrière Hospital, Paris), Pauline Dodet, MD (Pitié-Salpêtrière Hospital, Paris);

Genetic data: Jean-Christophe Corvol, MD, PhD (Pitié-Salpêtrière Hospital, Paris, coordination), Graziella Mangone, MD, PhD (Pitié-Salpêtrière Hospital, Paris, co-coordination), Samir Bekadar, MS (Pitié-Salpêtrière Hospital, Paris, biostatistician), Alexis Brice, MD (ICM, Pitié-Salpêtrière Hospital, Paris), Suzanne Lesage, PhD (INSERM, ICM, Paris, genetic analyses);

Metabolomics: Fanny Mochel, MD, PhD (Pitié-Salpêtrière Hospital, Paris, coordination), Farid Ichou, PhD (ICAN, Pitié-Salpêtrière Hospital, Paris), Vincent Perlbarg, PhD, Pierre and Marie Curie University), Benoit Colsch, PhD (CEA, Saclay), Arthur Tenenhaus, PhD (Supelec, Gif-sur-Yvette, data integration);

Brain MRI data: Stéphane Lehericy, MD, PhD (Pitié-Salpêtrière Hospital, Paris, coordination), Rahul Gaurav, MS, (Pitié-Salpêtrière Hospital, Paris, data analysis), Nadya Pyatigorskaya, MD, PhD, (Pitié-Salpêtrière Hospital, Paris, data analysis); Lydia Yahia-Cherif, PhD (ICM, Paris, Biostatistics), Romain Valabregue, PhD (ICM, Paris, data analysis), Cécile Galléa, PhD (ICM, Paris);

DaTscan imaging data: Marie-Odile Habert, MCU-PH (Pitié-Salpêtrière Hospital, Paris, coordination);

Voice recording: Dijana Petrovska, PhD (Telecom Sud Paris, Evry, coordination), Laetitia Jeancolas, MS (Telecom Sud Paris, Evry);

Study management: Vanessa Brochard (Pitié-Salpêtrière Hospital, Paris, coordination), Alizé Chalançon (Pitié-Salpêtrière Hospital, Paris, Project manager), Carole Dongmo-Kenfack (Pitié-Salpêtrière Hospital, Paris, clinical research assistant); Christelle Laganot (Pitié-Salpêtrière Hospital, Paris, clinical research assistant), Valentine Maheo (Pitié-Salpêtrière Hospital, Paris, clinical research assistant), Manon Gomes (Pitié-Salpêtrière Hospital, Paris, clinical research assistant).

## Authors’ contributions

SJ contributed to the design of the study, analyzed and interpretated the data, and wrote the first draft of the manuscript. SR conceived and designed the study, acquired, analyzed and interpreted the data, and substantially revised the manuscript. ADe, CT, AV, MFil, MTw, JPT, JTO, MFir, AT, PCD, JK, MH, PD, SM, ZV, SL, IA, MV, JCC, JFG, RBP, ADa, RC, HC, SL, EM, KAEM, LC, MS, SR, PB, KK, AKH, DA, BO, PM, LR, OM contributed to data acquisition and revision of the manuscript. All authors read and approved the final manuscript.

## References

1. Spillantini MG, Schmidt ML, Lee VM-Y, Trojanowski JQ, Jakes R, Goedert M. α-Synuclein in Lewy bodies. Nature. 1997;388:839–40.

2. Darweesh SKL, Verlinden VJA, Stricker BH, Hofman A, Koudstaal PJ, Ikram MA. Trajectories of prediagnostic functioning in Parkinson’s disease. Brain. 2017;140:429–41.

3. Berg D, Borghammer P, Fereshtehnejad SM, Heinzel S, Horsager J, Schaeffer E, et al. Prodromal Parkinson disease subtypes — key to understanding heterogeneity. Nature Reviews Neurology. 2021;17:349–61.

4. Joza S, Hu MT, Jung K-Y, Kunz D, Stefani A, Dušek P, et al. Progression of clinical markers in prodromal Parkinson’s disease and dementia with Lewy bodies: a multicentre study. Brain. 2023;awad072.

5. Rahayel S, Tremblay C, Vo A, Zheng YQ, Lehéricy S, Arnulf I, et al. Brain atrophy in prodromal synucleinopathy is shaped by structural connectivity and gene expression. Brain. 2022;145:3162–78.

6. Rahayel S, Tremblay C, Vo A, Misic B, Lehéricy S, Arnulf I, et al. Mitochondrial function-associated genes underlie cortical atrophy in prodromal synucleinopathies. Brain. 2023;awad044.

7. Rahayel S, Postuma RB, Montplaisir J, Genier Marchand D, Escudier F, Gaubert M, et al. Cortical and subcortical gray matter bases of cognitive deficits in REM sleep behavior disorder. Neurology. 2018;90:e1759–70.

8. Rahayel S, Postuma RB, Montplaisir J, Mišić B, Tremblay C, Vo A, et al. A Prodromal Brain-Clinical Pattern of Cognition in Synucleinopathies. Annals of Neurology. 2021;89:341–57.

9. Pereira JB, Weintraub D, Chahine L, Aarsland D, Hansson O, Westman E. Cortical thinning in patients with REM sleep behavior disorder is associated with clinical progression. npj Parkinson’s Disease 2019 5:1. 2019;5:1–8.

10. Joza S, Hu MT, Jung K-Y, Kunz D, Arnaldi D, Lee J-Y, et al. Prodromal dementia with Lewy bodies in REM sleep behavior disorder: A multicenter study. Alzheimer’s & Dementia. 2024;20:91–102.

11. Rahayel S, Mišić B, Zheng Y-Q, Liu Z-Q, Abdelgawad A, Abbasi N, et al. Differentially targeted seeding reveals unique pathological alpha-synuclein propagation patterns. Brain. 2021;145:1743–56.

12. Zheng Y-Q, Zhang Y, Yau Y, Zeighami Y, Larcher K, Misic B, et al. Local vulnerability and global connectivity jointly shape neurodegenerative disease propagation. PLoS Biol. 2019;17:e3000495.

13. Abdelgawad A, Rahayel S, Zheng Y-Q, Tremblay C, Vo A, Misic B, et al. Predicting longitudinal brain atrophy in Parkinson’s disease using a Susceptible-Infected-Removed agent-based model. Netw Neurosci. 2023;7:906–25.

14. Campabadal A, Segura B, Junque C, Iranzo A. Structural and functional magnetic resonance imaging in isolated REM sleep behavior disorder: A systematic review of studies using neuroimaging software. Sleep Med Rev. 2021;59:101495.

15. Shin JH, Kim H, Kim YK, Yoon EJ, Nam H, Jeon B, et al. Longitudinal evolution of cortical thickness signature reflecting Lewy body dementia in isolated REM sleep behavior disorder: a prospective cohort study. Transl Neurodegener. 2023;12:27.

16. Young AL, Marinescu RV, Oxtoby NP, Bocchetta M, Yong K, Firth NC, et al. Uncovering the heterogeneity and temporal complexity of neurodegenerative diseases with Subtype and Stage Inference. Nature Communications 2018 9:1. 2018;9:1–16.

17. Chertkow H, Borrie M, Whitehead V, Black SE, Feldman HH, Gauthier S, et al. The Comprehensive Assessment of Neurodegeneration and Dementia: Canadian Cohort Study. Can J Neurol Sci. 2019;46:499–511.

18. Duchesne S, Chouinard I, Potvin O, Fonov VS, Khademi A, Bartha R, et al. The Canadian Dementia Imaging Protocol: Harmonizing National Cohorts. J Magn Reson Imaging. 2019;49:456–65.

19. Mohaddes Z, Das S, Abou-Haidar R, Safi-Harab M, Blader D, Callegaro J, et al. National Neuroinformatics Framework for Canadian Consortium on Neurodegeneration in Aging (CCNA). Front Neuroinform [Internet]. 2018 [cited 2024 Jul 15];12. Available from: https://www.frontiersin.org/journals/neuroinformatics/articles/10.3389/fninf.2018.00085/full

20. Marek K, Chowdhury S, Siderowf A, Lasch S, Coffey CS, Caspell-Garcia C, et al. The Parkinson’s progression markers initiative (PPMI) - establishing a PD biomarker cohort. Ann Clin Transl Neurol. 2018;5:1460–77.

21. American Academy of Sleep Medicine, editor. International classification of sleep disorders. 3. ed. Darien, Ill: American Acad. of Sleep Medicine; 2014.

22. McKeith IG, Boeve BF, Dickson DW, Halliday G, Taylor J-P, Weintraub D, et al. Diagnosis and management of dementia with Lewy bodies. Neurology. 2017;89:88–100.

23. Stiasny-Kolster K, Mayer G, Schäfer S, Möller JC, Heinzel-Gutenbrunner M, Oertel WH. The REM sleep behavior disorder screening questionnaire--a new diagnostic instrument. Movement disorders : official journal of the Movement Disorder Society. 2007;22:2386–93.

24. Klapwijk ET, van de Kamp F, van der Meulen M, Peters S, Wierenga LM. Qoala-T: A supervised-learning tool for quality control of FreeSurfer segmented MRI data. NeuroImage. 2019;189:116–29.

25. Monereo-Sánchez J, de Jong JJA, Drenthen GS, Beran M, Backes WH, Stehouwer CDA, et al. Quality control strategies for brain MRI segmentation and parcellation: Practical approaches and recommendations - insights from the Maastricht study. NeuroImage. 2021;237:118174.

26. Rahayel S, Gaubert M, Postuma RB, Montplaisir J, Carrier J, Monchi O, et al. Brain atrophy in Parkinson’s disease with polysomnography-confirmed REM sleep behavior disorder. Sleep. 2019;42.

27. Voevodskaya O, Simmons A, Nordenskjöld R, Kullberg J, Ahlström H, Lind L, et al. The effects of intracranial volume adjustment approaches on multiple regional MRI volumes in healthy aging and Alzheimer’s disease. Front Aging Neurosci. 2014;6:264.

28. Vogel JW, Young AL, Oxtoby NP, Smith R, Ossenkoppele R, Strandberg OT, et al. Four distinct trajectories of tau deposition identified in Alzheimer’s disease. Nature Medicine. 2021;27:871–81.

29. Scotton WJ, Shand C, Todd E, Bocchetta M, Cash DM, VandeVrede L, et al. Uncovering spatiotemporal patterns of atrophy in progressive supranuclear palsy using unsupervised machine learning. Brain Communications. 2023;5:fcad048.

30. Fortin, Jean-Philippe. neuroCombat: Harmonization of multi-site imaging data with ComBat [Internet]. 2023. Available from: https://github.com/Jfortin1/neuroCombat_Rpackage

31. Fortin J-P, Cullen N, Sheline YI, Taylor WD, Aselcioglu I, Cook PA, et al. Harmonization of cortical thickness measurements across scanners and sites. NeuroImage. 2018;167:104–20.

32. Radua J, Vieta E, Shinohara R, Kochunov P, Quidé Y, Green MJ, et al. Increased power by harmonizing structural MRI site differences with the ComBat batch adjustment method in ENIGMA. Neuroimage. 2020;218:116956.

33. Aksman LM, Wijeratne PA, Oxtoby NP, Eshaghi A, Shand C, Altmann A, et al. pySuStaIn: A Python implementation of the Subtype and Stage Inference algorithm. SoftwareX. 2021;16:100811.

34. Guillerme T. dispRity: A modular R package for measuring disparity. Methods in Ecology and Evolution. 2018;9:1755–63.

35. Marinescu RV, Eshaghi A, Alexander DC, Golland P. BrainPainter: A software for the visualisation of brain structures, biomarkers and associated pathological processes. Multimodal Brain Image Analysis and Mathematical Foundations of Computational Anatomy : 4th International Workshop, MBIA 2019, and 7th International Workshop, MFCA 2019, held in conjunction with MICCAI 2019, Shenzhen, China, October 17,. 2019;11846:112.

36. Fasnacht JS, Wueest AS, Berres M, Thomann AE, Krumm S, Gutbrod K, et al. Conversion between the Montreal Cognitive Assessment and the Mini-Mental Status Examination. Journal of the American Geriatrics Society. 2023;71:869–79.

37. Campabadal A, Inguanzo A, Segura B, Serradell M, Abos A, Uribe C, et al. Cortical gray matter progression in idiopathic REM sleep behavior disorder and its relation to cognitive decline. NeuroImage: Clinical. 2020;28:102421.

38. Vo A, Tremblay C, Rahayel S, Shafiei G, Hansen JY, Yau Y, et al. Network connectivity and local transcriptomic vulnerability underpin cortical atrophy progression in Parkinson’s disease. Neuroimage Clin. 2023;40:103523.

39. Shafiei G, Bazinet V, Dadar M, Manera AL, Collins DL, Dagher A, et al. Network structure and transcriptomic vulnerability shape atrophy in frontotemporal dementia. Brain. 2023;146:321–36.

40. Kantarci K, Nedelska Z, Chen Q, Senjem ML, Schwarz CG, Gunter JL, et al. Longitudinal atrophy in prodromal dementia with Lewy bodies points to cholinergic degeneration. Brain Communications. 2022;4:fcac013.

41. Oppedal K, Ferreira D, Cavallin L, Lemstra AW, ten Kate M, Padovani A, et al. A signature pattern of cortical atrophy in dementia with Lewy bodies: A study on 333 patients from the European DLB consortium. Alzheimer’s & Dementia. 2019;15:400–9.

42. Constant AB, Basavaraju R, France J, Honig LS, Marder KS, Provenzano FA, et al. Longitudinal Patterns of Cortical Atrophy on MRI in Patients With Alzheimer Disease With and Without Lewy Body Pathology. Neurology. 2022;99:e1843–52.

43. Nedelska Z, Ferman TJ, Boeve BF, Przybelski SA, Lesnick TG, Murray ME, et al. Pattern of brain atrophy rates in autopsy-confirmed dementia with Lewy bodies. Neurobiology of Aging. 2015;36:452–61.

44. Zhou C, Wang L, Cheng W, Lv J, Guan X, Guo T, et al. Two distinct trajectories of clinical and neurodegeneration events identified in Parkinson’s disease [Internet]. In Review; 2022 Aug. Available from: https://www.researchsquare.com/article/rs-1880346/v1

45. Fereshtehnejad SM, Yao C, Pelletier A, Montplaisir JY, Gagnon JF, Postuma RB. Evolution of prodromal Parkinson’s disease and dementia with Lewy bodies: a prospective study. Brain. 2019;142:2051–67.

46. Postuma RB, Iranzo A, Hu M, Högl B, Boeve BF, Manni R, et al. Risk and predictors of dementia and parkinsonism in idiopathic REM sleep behaviour disorder: a multicentre study. Brain. 2019;142:744–59.

47. Sarro L, Senjem ML, Lundt ES, Przybelski SA, Lesnick TG, Graff-Radford J, et al. Amyloid-β deposition and regional grey matter atrophy rates in dementia with Lewy bodies. Brain. 2016;139:2740–50.

48. Irwin DJ, Hurtig HI. The Contribution of Tau, Amyloid-Beta and Alpha-Synuclein Pathology to Dementia in Lewy Body Disorders. J Alzheimers Dis Parkinsonism. 2018;8:444.

49. Fernandes M, Maio S, Eusebi P, Placidi F, Izzi F, Spanetta M, et al. Cerebrospinal-fluid biomarkers for predicting phenoconversion in patients with isolated rapid-eye movement sleep behavior disorder. Sleep. 2024;47:zsad198.

50. Diaz-Galvan P, Przybelski SA, Lesnick TG, Schwarz CG, Senjem ML, Gunter JL, et al. β-Amyloid Load on PET Along the Continuum of Dementia With Lewy Bodies. Neurology. 2023;101:e178–88.

51. Borghammer P. The brain-first vs. body-first model of Parkinson’s disease with comparison to alternative models. J Neural Transm (Vienna). 2023;130:737–53.

52. Marchand DG, Postuma RB, Escudier F, Roy JD, Pelletier A, Montplaisir J, et al. How does dementia with Lewy bodies start? prodromal cognitive changes in REM sleep behavior disorder. Annals of Neurology. 2018;83:1016–26.

53. Borghammer P, Okkels N, Weintraub D. Parkinson’s Disease and Dementia with Lewy Bodies: One and the Same. JPD. 2024;14:383–97.

54. Torontali ZA, Fraigne JJ, Sanghera P, Horner R, Peever J. The Sublaterodorsal Tegmental Nucleus Functions to Couple Brain State and Motor Activity during REM Sleep and Wakefulness. Current Biology. 2019;29:3803–3813.e5.

55. Biondetti E, Santin MD, Valabrègue R, Mangone G, Gaurav R, Pyatigorskaya N, et al. The spatiotemporal changes in dopamine, neuromelanin and iron characterizing Parkinson’s disease. Brain. 2021;144:3114–25.

56. Nepozitek J, Varga Z, Dostalova S, Perinova P, Keller J, Robinson S, et al. Magnetic susceptibility changes in the brainstem reflect REM sleep without atonia severity in isolated REM sleep behavior disorder. npj Parkinsons Dis. 2023;9:1–8.

