## Supplementary for "Distinct brain atrophy progression subtypes underlie phenoconversion in isolated REM sleep behaviour disorder"

### Data Supplement:

**Supplementary Table 1: Sample size and MRI acquisition parameters by study centre**

| Centre | Controls | iRBD | DLB | PD-pRBD | Scanner | Sequence | TR (ms) | TE (ms) | Flip angle | Voxel size |
|---|---|---|---|---|---|---|---|---|---|---|
| Aarhus | 19/20 | 13/18 | - | - | 3T Siemens MAGNETOM Skyra 32c head coil | MPRAGE | 2420 | 3.7 | 9° | 1 mm³ isotropic |
| CCNA | 57/57 | - | 13/14 | - | See reference 18 | - | - | - | - | - |
| Cologne | - | 31/47 | - | - | 1.5T Philips INGENIA 16c head coil | MPRAGE | 7.6 | 3.5 | 8° | 0.55 x 0.55 x 0.95 mm |
| Genoa | 13/15 | 10/14 | - | - | 3T Siemens PRISMA 64c head coil | MPRAGE | 2300 | 2.98 | 9° | 1 mm³ isotropic |
| Montreal | 84/94 | 73/84 | - | - | 3T Siemens TIM Trio 12c head coil | MPRAGE | 2300 | 2.91 | 9° | 1 mm³ isotropic |
| | | | | | 3T Siemens MAGNETOM Prisma 32c head coil | MPRAGE | 2300 | 2.98 | 9° | 1 mm³ isotropic |
| Newcastle | 59/62 | - | 69/73 | - | 3T Philips Achieva 8c head coil | MPRAGE | 8.3 | 4.6 | 8° | 1 mm³ isotropic |
| | | | | | 3T Philips Achieva, 8c head coil | MPRAGE | 9.6 | 4.6 | 8° | 1.15 x 1.15 x 1.2 mm |
| Oxford | 59/66 | 73/81 | - | - | 3T Siemens Trio 12c head coil | MPRAGE | 2040 | 4.7 | 8° | 1 mm³ isotropic |
| PPMI | 92/126 | 17/37 | - | 110/142 | See reference 17 | - | - | - | - | - |
| Paris | 60/73 | 49/57 | - | - | 3T Siemens TIM Trio 12c head coil | MPRAGE | 2300 | 4.18 | 9° | 1 mm³ isotropic |
| | | | | | 3T PRISMA Fit 64c head coil | MPRAGE2 | 5000 | 2.98 | 4° & 5° | 1 mm³ isotropic |
| Prague | 55/57 | 75/83 | - | - | 3T Siemens SKYRA 32c head coil | MPRAGE | 2200 | 2.4 | 8° | 1 mm³ isotropic |
| Sydney | 21/26 | 21/30 | - | - | 3T GE Discovery MR750 8c head coil | BRAVO | 5800 | 2.6 | 12° | 1 mm³ isotropic |
| Total | 519/596 | 362/451 | 82/87 | 110/142 | 3T Siemens MAGNETOM Skyra 32c head coil | MPRAGE | 2420 | 3.7 | 9° | 1 mm³ isotropic |

Sample sizes reflect the number of patients used in the SuStaIn modelling. The first number reflects the number of patients who passed quality control, whereas the second number reflects the total number of patients imaged.

CCNA = Canadian Consortium on Neurodegeneration in Aging; DLB = dementia with Lewy bodies; iRBD = idiopathic/isolated REM sleep behaviour disorder; PD-pRBD = Parkinson's disease with possible REM sleep behaviour disorder; PPMI = Parkinson's Progression Markers Initiative.

**Supplementary Table 2: Descriptive statistics of patients by group**

| Variable | Controls | iRBD | DLB | PD-pRBD | p-value | Post-hoc testing |
|---|---|---|---|---|---|---|
| Age | 65.6 ± 10.1 | 67.1 ± 6.95 | 76.8 ± 6.45 | 62.1± 8.93 | **<0.001** | DLB > Controls = iRBD > PD |
| Sex (% female) | 41.4 | 13.3 | 29.3 | 28.7 | **<0.001** | Control = DLB = PD > iRBD |
| MoCA | 26.8 ± 2.36 | 25.7 ± 3.02 | 14.4 ± 5.46 | 25.9 ± 3.31 | **<0.001** | Controls = PD > iRBD > DLB |
| MDS-UPDRS-III | 2.28 ± 4.44 | 6.04 ± 5.57 | 32.1 ± 18.1 | 21.4 ± 9.61 | **<0.001** | DLB > PD > iRBD > Controls |

Groups were compared using ANOVA with Tukey HSD post-hoc tests for continuous variables. Chi-squared tests were used with post-hoc pairwise testing for categorical variables.

DLB = dementia with Lewy bodies; iRBD = idiopathic/isolated REM sleep behaviour disorder; MoCA = Montreal Cognitive Assessment; MDS-UPDRS-III = Movement Disorders Society – Unified Parkinson's Disease Rating Scale, Part III; PD-pRBD = Parkinson's disease with possible REM sleep behaviour disorder; SD = standard deviation.

**Supplementary Table 3 Clinical variables associated to each subtype when using PD-pRBD, iRBD, and DLB**

| | Classifiable | | | Subtyped | | |
|---|---|---|---|---|---|---|
| **Phenoconversion** | **Non-classifiable** | **Classifiable** | **p-value[a]** | **Cortical-first** | **Subcortical-first** | **p-value[b]** |
| **Demographics** | | | | | | |
| n (%): iRBD | 176 (70.4) | 186 (61.2) | | 111 (62.7) | 75 (59.1) | |
| n (%): DLB | 19 (7.6) | 63 (20.7) | | 33 (18.6) | 30 (23.6) | |
| n (%): PD-pRBD | 55 (22) | 55 (18.1) | | 33 (18.6) | 22 (17.3) | |
| Age: All | 67.1 (8.2) | 68 (8.7) | 0.218 | 68.1 (8.8) | 67.7 (8.7) | 0.654 |
| Age: iRBD | 67.3 (7.3) | 67 (6.6) | 0.667 | 67.3 (6.4) | 66.5 (7.1) | 0.472 |
| Age: DLB | 75.9 (6) | 77.1 (6.6) | 0.48 | 78.3 (6.9) | 75.7 (6) | 0.116 |
| Age: PD-pRBD | 63.3 (9) | 60.9 (8.8) | 0.151 | 61 (8.9) | 60.7 (8.8) | 0.921 |
| % male | 83.8 | 83.8 | 0.960 | 82.7 | 85.6 | 0.611 |
| Stage[c] (SD): All | 0 (0) | 4.3 (3.9) | **<0.001** | 4.0 (3.8) | 4.6 (3.9) | 0.232 |
| Stage[c] (SD): iRBD | 0 (0) | 3.6 (2.8) | **<0.001** | 3.2 (1.9) | 4.2 (3.6) | **0.029** |
| Stage[c] (SD): DLB | 0 (0) | 6.7 (6) | **<0.001** | 7.5 (7) | 5.7 (4.6) | 0.215 |
| Stage[c] (SD): PD-pRBD | 0 (0) | 3.8 (2.8) | **<0.001** | 3.4 (2) | 4.3 (3.7) | 0.293 |
| **Clinical variables** | | | | | | |
| MDS-UPDRS-III (SD): All | 10.4 (11.1) | 15.4 (15.2) | **<0.001** | 15.8 (15.9) | 14.8 (14.3) | 0.56 |
| MDS-UPDRS-III (SD): iRBD | 5.2 (4.9) | 6.9 (6.1) | **0.005** | 7.1 (6.6) | 6.5 (5.4) | 0.475 |
| MDS-UPDRS-III (SD): DLB | 24.4 (17.4) | 34.4 (17.8) | **0.036** | 37.4 (18.9) | 31.2 (16.2) | 0.166 |
| MDS-UPDRS-III (SD): PD-pRBD | 21.7 (9.8) | 21.1 (9.4) | 0.737 | 21.6 (9.2) | 20.4 (9.9) | 0.634 |
| MoCA (SD): All | 25.4 (3.7) | 23.3 (6) | **<0.001** | 23.3 (6.3) | 23.3 (5.6) | 0.984 |
| MoCA (SD): iRBD | 26.1 (2.7) | 25.4 (3.2) | **0.044** | 25.4 (3.4) | 25.4 (3) | 0.921 |
| MoCA (SD): DLB | 17.5 (5.2) | 13.5 (5.3) | **0.015** | 12.6 (5.9) | 14.6 (4.2) | 0.152 |
| MoCA (SD): PD-pRBD | 25.7 (3.1) | 26 (3.5) | 0.675 | 25.9 (3.4) | 26.1 (3.8) | 0.836 |
| % MCI: iRBD[d] | 36.3 | 45.3 | 0.084 | 43.0 | 48.6 | 0.452 |
| % MCI: PD-pRBD[d] | 40.0 | 36.7 | 0.745 | 37.9 | 35.0 | 0.834 |

Statistical differences were calculated using unpaired t-tests for continuous variables and chi-squared test for categorical variables.
[a]Non-classifiable group versus classifiable group.
[b]Cortical-first subtype versus subcortical-first subtype.
[c]Stage refers to SuStaIn stage.
[d]MCI as defined by ≤ 25/30 on MoCA; all DLB patients met criteria for dementia.

DLB = dementia with Lewy bodies; iRBD = idiopathic/isolated REM sleep behaviour disorder; MoCA = Montreal Cognitive Assessment; MCI = mild cognitive impairment; MDS-UPDRS-III = Movement Disorders Society – Unified Parkinson's Disease Rating Scale, Part III; MoCA = Montreal Cognitive Assessment; PD-pRBD = Parkinson's disease with possible REM sleep behaviour disorder; SD = standard deviation; SuStaIn = Subtype and Staging Inference.

**Supplementary Table 4 Phenoconversion outcomes in iRBD patients by SuStaIn subtypes**

| Phenoconversion | Classifiable | | | Subtyped | | |
|---|---|---|---|---|---|---|
| | Non-classifiable | Classifiable | p-value[a] | Cortical-first | Subcortical-first | p-value[b] |
| All converters | 41 | 43 | 0.904 | 23 | 10 | 0.311 |
| DLB converters | 15 | 13 | - | 8 | 5 | - |
| PD converters | 23 | 29 | - | 15 | 14 | - |
| MSA converters | 3 | 1 | - | 0 | 1 | - |

Statistical differences were calculated using chi-squared tests.
[a]Non-classifiable group versus classifiable group.
[b]Cortical-first subtype versus subcortical-first subtype.

DLB = dementia with Lewy bodies; iRBD = idiopathic/isolated REM sleep behaviour disorder; MSA = multiple system atrophy; PD = Parkinson's disease; SuStaIn = Subtype and Staging Inference.

**Supplementary Table 5 Risk of phenoconversion in iRBD based on SuStain classifiability**

| Variable | Estimate | Standard error | z-value | p-value |
|---|---|---|---|---|
| Classifiability | 3.383 | 1.646 | 2.056 | **0.040** |
| Age | 0.000 | 0.019 | -0.022 | 0.983 |
| Sex | -0.267 | 0.392 | -0.680 | 0.497 |
| Classifiability*probability of classifiability | -4.318 | 2.032 | -2.125 | **0.034** |

Logistic regression model of phenoconversion ~ age + sex + classifiability * probability of classifiability
Classifiability is coded as 0 = non-classifiable and 1 = classifiable as either cortical-first or subcortical-first subtypes

iRBD = idiopathic/isolated REM sleep behaviour disorder; SuStaIn = Subtype and Staging Inference.

**Supplementary Table 6 Risk of phenoconversion in iRBD based on the SuStain solution**

| Variable | Estimate | Standard error | z-value | p-value |
|---|---|---|---|---|
| Subtype | 1.449 | 0.634 | 2.286 | **0.022** |
| Stage | 0.390 | 0.242 | 1.613 | 0.107 |
| Age | 0.038 | 0.030 | 1.294 | 0.196 |
| Sex | 0.177 | 0.560 | 0.316 | 0.752 |
| Subtype * Stage | -0.313 | 0.159 | -1.973 | **0.049** |

Logistic regression model of phenoconversion ~ age + sex + subtype * stage.
Subtype is coded as 0 = cortical-first subtype and 1 = subcortical-first subtype.
Stage refers to SuStaIn stage.

iRBD = idiopathic/isolated REM sleep behaviour disorder; SuStaIn = Subtype and Staging Inference.

**Supplementary Table 7 Risk of phenoconversion in iRBD by SuStaIn subtypes**

| Variable | Estimate | Standard error | z-value | p-value |
|---|---|---|---|---|
| **Subcortical-first subtype** | | | | |
| Stage | -0.232 | 0.120 | -1.930 | **0.054** |
| Age | -0.014 | 0.044 | -0.323 | 0.746 |
| Sex | -0.129 | 0.825 | -0.157 | 0.875 |
| **Cortical-first subtype** | | | | |
| Stage | 0.054 | 0.108 | 0.502 | 0.615 |
| Age | 0.082 | 0.042 | 1.966 | **0.049** |
| Sex | 0.494 | 0.765 | 0.646 | 0.519 |

Logistic regression model of phenoconversion ~ stage + subtype + age + sex.
Stage refers to SuStaIn stage.

iRBD = idiopathic/isolated REM sleep behaviour disorder; SuStaIn = Subtype and Staging Inference.

**Supplementary Table 8 Risk of specific phenoconversion in iRBD by SuStaIn subtypes**

| Variable | Estimate | Standard error | z-value | p-value |
|---|---|---|---|---|
| **Parkinsonism-predominant subtype** | | | | |
| Subtype | 0.632 | 0.257 | 2.455 | **0.014** |
| Stage | -0.270 | 0.213 | -1.268 | 0.205 |
| Age | -0.246 | 0.479 | -0.513 | 0.608 |
| Sex | -0.001 | 0.022 | -0.043 | 0.966 |
| Subtype * Stage | 0.037 | 0.132 | 0.278 | 0.781 |
| **Dementia-predominant subtype** | | | | |
| Subtype | -0.121 | 0.407 | -0.298 | 0.766 |
| Stage | 0.366 | 0.190 | 1.923 | **0.055** |
| Age | 0.028 | 0.031 | 0.906 | 0.365 |
| Sex | -0.081 | 0.647 | -0.125 | 0.901 |
| Subtype * Stage | -0.197 | 0.146 | -1.350 | 0.177 |

Separate logistic regression models of parkinsonism (or dementia) ~ age + sex + subtype * stage.
Subtype is coded as 0 = cortical-first subtype and 1 = subcortical first subtype.
Stage refers to SuStaIn stage.

iRBD = idiopathic/isolated REM sleep behaviour disorder; SuStaIn = Subtype and Staging Inference.

**Supplementary Figure 1: SuStaIn modelling using cortical volume and subcortical volume**

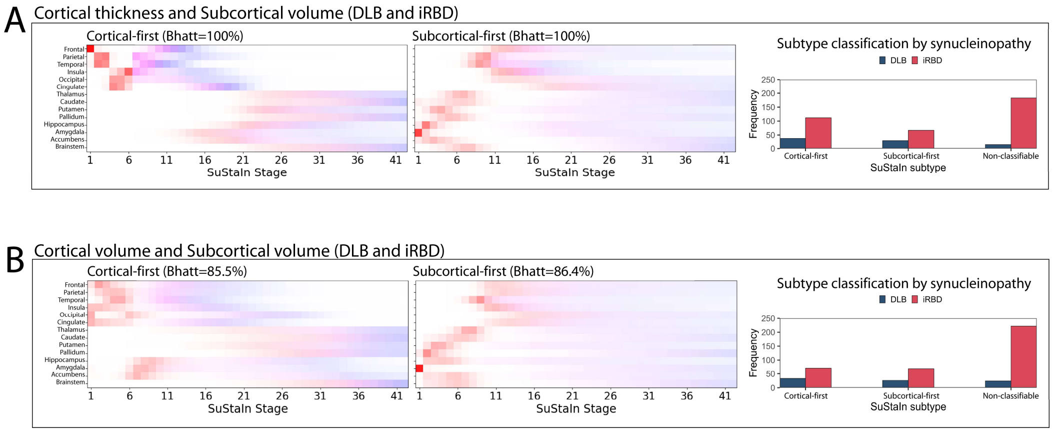


Positional variance diagram demonstrating two SuStaIn subtypes using **(A)** cortical thickness + subcortical volumes or **(B)** cortical volume + subcortical volumes as inputs into the model. In both models, SuStaIn identified two unique subtypes of brain atrophy progression with similar patterns of progression, as expressed by the Bhattacharyya coefficient. At each stage, the colour in each region indicates the level of severity of atrophy, with white representing unaffected regions, red mildly affected regions (z-score of -1), magenta moderately affected regions (z-score of -2), and blue severely affected regions (z-score of -3 or more). The number of patients that were classified into a subtype or remained unclassified are shown in the graphs on the right.

Bhatt = Bhattacharyya coefficient; DLB = dementia with Lewy bodies; iRBD = idiopathic/isolated REM sleep behaviour disorder; SuStaIn = Subtype and Staging Inference.

**Supplementary Figure 2: Effect of adding different synucleinopathies on the SuStaIn solution**

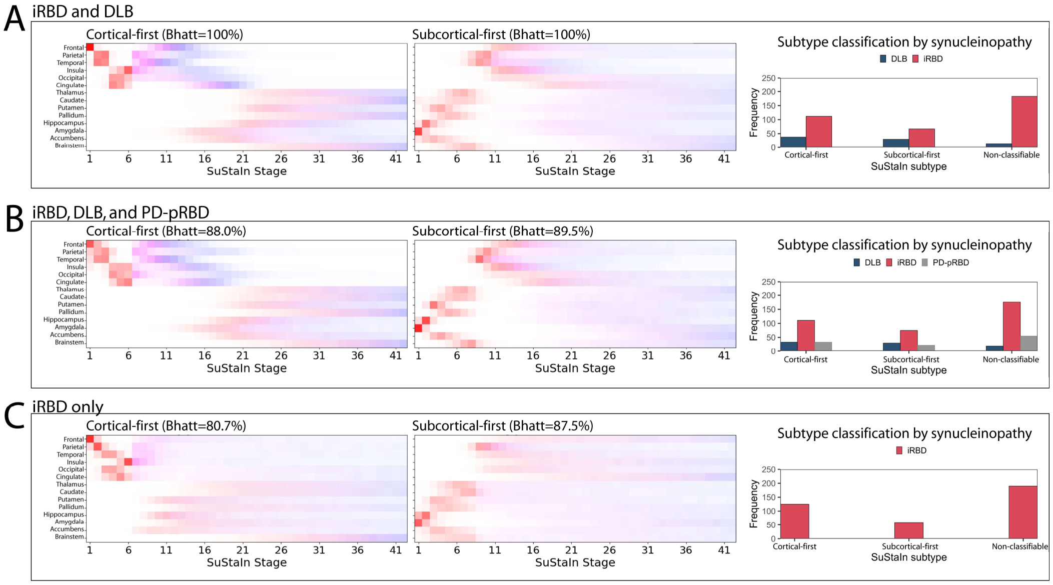


Positional variance diagrams comparing SuStaIn modelling using as input patients the **(A)** iRBD and DLB patients only – the primary model, **(B)** iRBD, DLB, and PD-pRBD patients, and **(C)** iRBD patients only. In each case, SuStaIn identified two unique subtypes of brain atrophy progression with similar patterns of progression as determined by the Bhattacharyya coefficient when comparing models (B) and (C) to the primary SuStaIn model used (A). At each stage, the colour in each region indicates the level of severity of atrophy, where white represents unaffected regions, red mildly affected regions (z-score of -1), magenta moderately affected regions (z-score of -2), and blue severely affected regions (z-score of -3 or less). Increased uncertainty of staging is observed at higher SuStaIn stages when DLB patients are not included in the modelling ("smudging"), resulting in a lower Bhattacharyya coefficient. The number of patients that were classified into a subtype or remained unclassified are shown in the graphs on the right.

Bhatt = Bhattacharyya coefficient; DLB = dementia with Lewy bodies; iRBD = idiopathic/isolated REM sleep behaviour disorder; PD-pRBD = Parkinson's disease with probable REM sleep behaviour disorder; SuStaIn = Subtype and Staging Inference.
